# Gene-by-Sleep Duration Interaction for Glycemic Traits in over 480,000 Individuals

**DOI:** 10.64898/2026.03.02.26346498

**Authors:** Heming Wang, Pavithra Nagarajan, Clint L Miller, Amy R Bentley, Raymond Noordam, Kenneth E Westerman, Michael R Brown, Aldi T Kraja, Jeffrey R O’Connell, Karen Schwander, Changwei Li, Mihir M Sanghvi, Yipei Song, Traci M Bartz, Vincent Braunack-Mayer, Ling Chen, Jiawen Du, Diana Dunca, Mary F Feitosa, Valborg Gudmundsdottir, Xiuqing Guo, Sarah E Harris, Heather M Highland, Zhijie Huang, Changhoon Kang, Timo A Lakka, Christophe Lefevre, Jian’an Luan, Leo-Pekka Lyytikäinen, Celestin Missikpode, John L Morrison, Nicholette D Palmer, Anne Richmond, Mina Shahisavandi, Jingxian Tang, Peter J van der Most, Stefan Weiss, Chenglong Yu, Wanying Zhu, Md Abu Yusuf Ansari, Pramod Anugu, Hugues Aschard, Kaavya Ashok, John R Attia, Lydia A Bazzano, Brian E Cade, Archie Campbell, Latchezar M Dimitrov, Anh Do, Tariq Faquih, Sandy L FinesilverSmith, Susan P Fisher-Hoch, Amanda M Fretts, Sina A Gharib, Mark O Goodarzi, Mariaelisa Graff, Charles Gu, Paul Hanson, Jiang He, Sami Heikkinen, James Hixson, Sarah Hsu, Mika Kähönen, Minjung Kho, Hyunju Kim, Pirjo Komulainen, Lenore J Launer, Rozenn N Lemaitre, Lifelines Cohort Study, John J McNeil, Joseph B McCormick, Ilja M Nolte, Laura M Raffield, Olli T Raitakari, Julia Ramírez, Renata L Riha, Martin Risch, Lorenz Risch, Tom C Russ, Chloé Sarnowski, Miranda T Schram, Rodney J Scott, Tamar Sofer, Quan Sun, Uwe Völker, Henry Völzke, Yujie Wang, Ko Willems van Dijk, Alexis C Wood, Kristin L Young, Ruiyuan Zhang, Xiaofeng Zhu, Jennifer E Below, David Conen, Simon R Cox, Ervin R Fox, Nora Franceschini, Mohsen Ghanbari, Hans Jörgen Grabe, Vilmundur Gudnason, Caroline Hayward, Elizabeth G Holliday, Cashell E Jaquish, Paul Lacaze, Seunggeun Lee, Terho Lehtimäki, Ching-Ti Liu, Alanna C Morrison, Kari E North, Patricia A Peyser, Michael A Province, Bruce M Psaty, Rainer Rauramaa, Frits R Rosendaal, Jerome I Rotter, Harold Snieder, Lynne E Wagenknecht, Nicholas J Wareham, Ayush Giri, Tanika N Kelly, Patricia B Munroe, James Gauderman, Thomas W Winkler, Paul S de Vries, Dabeeru C Rao, Alisa K Manning, Han Chen, Lisa de las Fuentes, Susan Redline, James B Meigs

## Abstract

Both short and long sleep duration have been associated with poor glycemic control and an increased risk of developing type 2 diabetes mellitus. Although sleep duration may differentially modify the effects of genetic risk factors for type 2 diabetes, this has not been systematically investigated. In the present study, we conducted genome-wide gene by sleep duration meta-analyses, separately assessing interactions of short and long sleep, for fasting glucose, fasting insulin, and hemoglobin A1c in up to 489,309 individuals without diabetes from seven different population groups. In total, 16 loci were identified to interact with sleep duration — six with short sleep and ten with long sleep. Of these, four loci were identified through cross-population meta-analysis. Mapped genes exhibit pathway connections to pericyte apoptosis, NMDA receptor activity, the GLUT1 receptor, neurological health, and sleep architecture. Eleven loci (*VRK2, PCDH7, TFAP2A, CAP2, PAPPA, ZCCHC2, MYH9, SGIP1, JAKMIP3, RRAS2, MAPT*) have not been reported in previous glycemic trait genome-wide association studies. Interaction loci identify divergent biological mechanisms for short and long sleep duration influencing glycemic control, suggesting specific pathways of intervention for precision medicine approaches to diabetes prevention and management.

**Article Highlights:**

- The biological mechanisms of how sleep duration impacts type 2 diabetes pathogenesis and glycemic control are unclear.
- This study reveals 16 loci (11 novel) that interact with either short or long sleep duration to influence hemoglobin A1c, fasting glucose, or fasting insulin. Short and long sleep duration loci were non-overlapping.
- Regulation of copper and diacylglycerol levels appear as distinct cellular mechanisms implicated by long and short sleep duration loci respectively.
- Identified gene targets present insight for potential type 2 diabetes therapeutic design approaches related to JIP1-JNK interaction disruption, pericyte health, NMDA receptor activity, anti-inflammatory and leptin-enhancing dietary supplements, and serpins.

## Introduction

Type 2 diabetes is a disorder with an immense global health burden reflected by its attributable mortality projected to reach 1.63 million deaths by 2030 (1). It is also highly heterogeneous, with genetic studies undertaken to disentangle its endophenotypes revealing multiple biological axes for consideration, including vascular health and lipodystrophy (2). Genome-wide association analysis (GWAS) of glycemic traits in individuals without type 2 diabetes have identified over 200 genetic loci, revealing the importance of varying pathways ranging from the circadian clock, and lipid biology, to hematopoiesis (3).

Sleep disturbances are associated with type 2 diabetes risk (4). Yet, their specific roles in pathways underlying type 2 diabetes onset are unclear. Short sleep duration may disrupt cortisol rhythm, elevate lipolysis and sympathetic nervous system activity, and lower cerebral glucose utilization (5; 6). Long sleep duration may be linked to chronic inflammation and the misalignment of circadian rhythm within 24 hours (7). In particular, circadian rhythm plays an important role in glycemic regulation, demonstrated by rhythmicity of glycogen expression in liver and muscle, and strong diurnal variation of both blood glucose and insulin levels (8). With genomics able to offer a granular view of biological mechanisms, it is critical to investigate the relationship between sleep lifestyle and genetic variants underlying glycemic control.

We hypothesize that short and long sleep duration may distinctly modify genetic risk factor effects for glycemic control. Our prior genome-wide gene-by-sleep duration interaction analyses within the CHARGE Gene-Lifestyle Interaction Working Group have identified variant-sleep interactions, leading to the discovery of novel loci for blood pressure and lipid traits (9; 10). In this study, we investigate gene-by-short and gene-by-long sleep interactions for three glycemic traits which are instrumental to type 2 diabetes pathophysiology: hemoglobin A1c (HbA1c), fasting glucose (GL), and fasting insulin (INS). To prioritize holistic insights, we incorporate a diverse sample across seven different population groups in up to 489,309 individuals without diabetes.

## Research Design and Methods

This work was approved by the Institutional Review Board of Washington University in St. Louis and complies with all relevant ethical regulations. For each of the participating cohorts, the appropriate ethics review board approved the data collection, and all participants provided informed consent.

### Data Harmonization

The overall analysis flow is provided in Figure 1A. Meta-analysis of genome-wide gene-sleep duration interactions for fasting GL (mg/dL), HbA1c (%), and fasting INS (µU/mL) were conducted in 52 population group-specific cohorts derived from 30 studies and 7 population groups — African (AFR; N: 14,477), Admixed American (AMR; N: 688), East Asian (EAS; N: 41,201), European (EUR; N: 413,152), Hispanic/Latinos (HIS; N: 13,164), Middle Eastern (MID; N: 1,062), and South Asian (SAS; N: 5,115) (Supplementary Tables 1-2). Samples were mapped to each population group by individual study teams, based on self-reported identity or genetic similarity to reference populations from the 1000 Genomes Phase 3 and the Human Genome Diversity Project (11). Additional details are provided in Supplementary Methods. Analysis was restricted to adults (≥ 18 years) with available data on at least one of the three glycemic traits, self-reported total sleep time (TST), and genomics. Individuals with diabetes or reporting TST <3 or >14 hours were excluded. HbA1c, Fasting GL, and Fasting INS were analyzed in cohort studies while HbA1c and GL – in fasted and non-fasting groups separately – were analyzed in biobank studies (Supplementary Methods). GL and INS were harmonized to standard units (1 mmol/L = 18 mg/dL and 1 pmol/L = 0.16 μU/mL). INS was natural logarithm transformed and adjusted for BMI by regressing ln(INS) on BMI based on prior GWAS (3). Glycemic trait outliers (> ±6 standard deviations or SD) were winsorized to ±6 SD. Short and long TST (STST, LTST) were defined as under 20^th^ and above 80^th^ study-specific percentiles of age, sex, and age×sex adjusted TST respectively (9).

**Figure 1:**
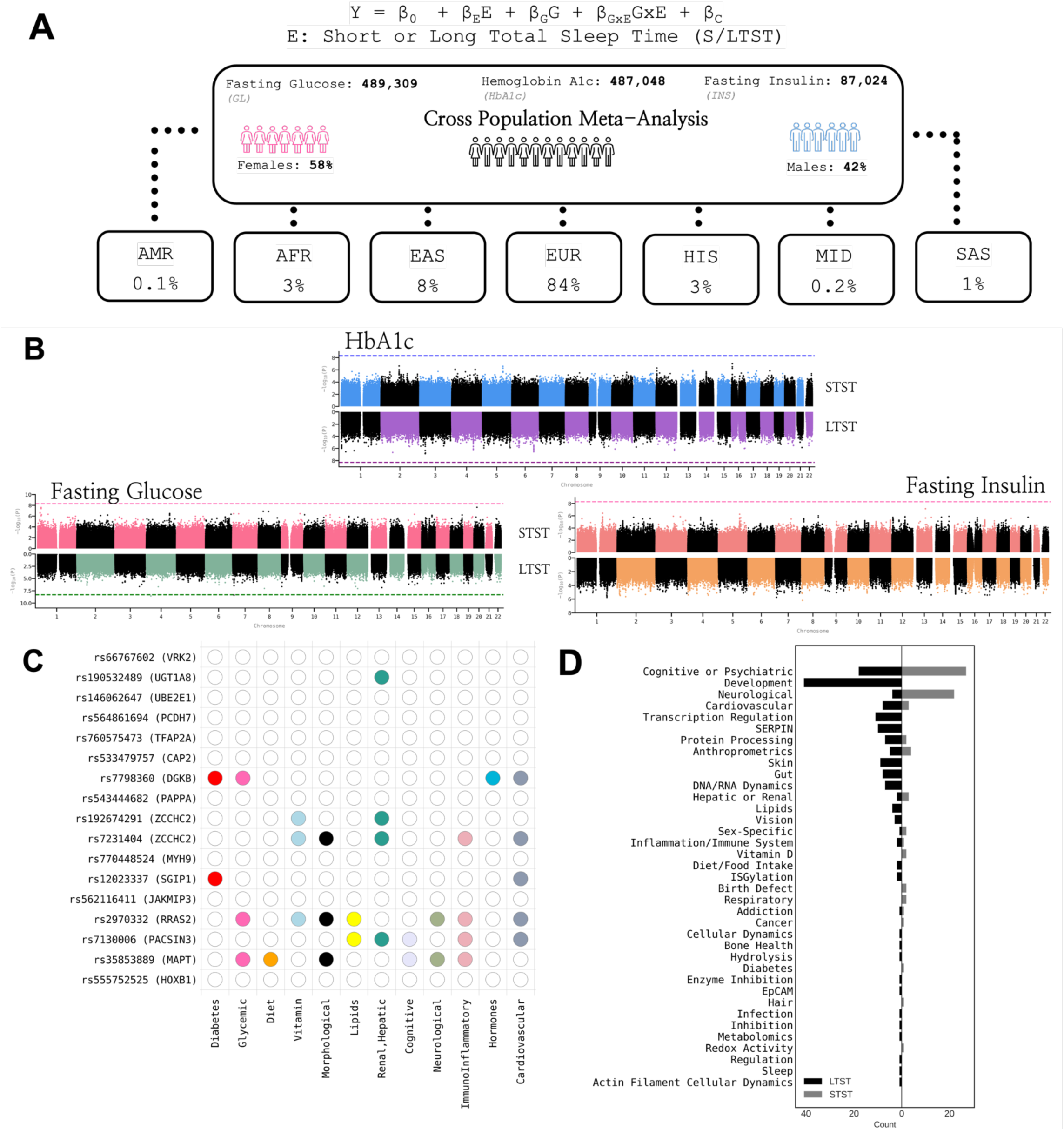
Gene-sleep duration interactions for glycemic traits. **A.** Analysis workflow. **B.** Miami plots of the 1df G×E interaction test for each of the 3 glycemic traits from combined sex cross-population meta analysis (CPMA), with the upper section of each plot denoting interaction with short total sleep time (STST), and lower section of each plot denoting interaction with long TST (LTST). **C**. Pleiotropic connections reported for each of the 17 lead variants discovered in this study queried across multiple GWAS databases (Supplementary Table 7). An empty cell denotes no significant (p<5e-08) trait connections, and a colored cell denotes presence of 1+ significant trait association(s). **D.** Gene set enrichment analysis results (FDR<0.05) across GENE2FUNC and STRING, via a horizontal bar plot displaying trait categories, grouped according to sleep exposure (STST: grey, LTST: black).

### Genome-Wide Gene-Environment Interaction Study (GWIS)

Each cohort or biobank study performed genome-wide analysis on variants with minor allele frequency (MAF)>0.1%, separately for each population group in combined sex, females and males. The primary analysis conducted was for the interaction model M1: *Y = β_M1_0_ + β_M1_E_E + β_M1_G_G + β_M1_GxE_ G× E + **β******_M1_C1_C_1_***, where Y denotes the glycemic trait, E denotes STST or LTST, G is a genetic variant, and ***C_1_*** is the set of covariates comprising of age, age^2^, sex, age×E, age^2^×E, sex×E, ancestry principal components (PCs), and study-specific covariates (e.g., study center). A secondary analysis using the marginal effect model M2: *Y = β_M2_0_ + β_M2_G_G + **β******_M2_C2_C_2_*** was conducted where ***C_2_*** included age, age^2^, sex, genomic PCs, and study-specific covariates.

### Meta-Analysis

Meta-analyses were subsequently conducted on the study-level summary statistics across and within each population and sex group. Fixed-effect inverse variance analysis was performed for 1 degree-of-freedom (df) tests of *P_M1_G,_ P_M1_GxE,_ P_M2_G,_* and the 2df test of *P_M1_G,GxE_* (12; 13). Genomic control correction was conducted before and after meta-analysis. Variants exhibiting significant STST or LTST interaction were identified by: 1) *P_M1_GxE_*<5×10⁻⁹ for the 1df test of the interaction, 2) *P_M1_G,GxE_* < 5×10⁻⁹, *P_M1_GxE_* < *P_M1_G_*, *P_M2_G_*>5×10⁻^8^ for the 2df joint test, or 3) a two-step method identifying a subset of variants (z) with *P_M2_G_*<1×10^-5^ that exhibit significant interaction *P_M1_GxE_* < 0.05/2z* where z* refers to the effective number of tests calculated from z (14). A false discovery rate (FDR) <0.05 threshold was additionally enforced. Lead variants were followed up for secondary bioinformatics analyses, including variant annotation, gene mapping, gene-set enrichment, and druggability analysis (Supplementary Methods).

### Data and Resource Availability

Genome-wide summary statistics of the cross-population gene-sleep duration interaction analyses are available in the GWAS Catalog. Source code for primary software used to conduct GxE meta-analysis is publicly available at the following repositories: GEM (https://github.com/large-scale-gxe-methods/GEM), MMAP (https://mmap.github.io/), LinGxEScanR (https://github.com/USCbiostats/LinGxEScanR), FUMA (https://fuma.ctglab.nl/), METAL (https://github.com/statgen/METAL).

## Results

### Overview

The total sample sizes included in this study were 489,309 for GL (58% female; 42% male); 487,048 for HbA1c (56% female; 44% male); and 87,024 for INS (68% female; 32% male) (Supplementary Table 2). In total, 17 lead variants mapped to 16 unique genomic loci (+/- 500 kilobases), and had a significant interaction with either short or long sleep duration (STST: 6, LTST: 11) across GL (10), HbA1c (4), and INS (3) (Table 1, Supplementary Table 3, Supplementary Figures 1-5). Seven, four, and six lead variants were identified by the 1df interaction test, the two-step method, and the 2df joint test, respectively. Additional loci with a more lenient threshold of *P_M1_G_*_×_*_E_* or *P_M1_G,G_*_×_*_E_* < 5×10^-8^ are summarized in Supplementary Tables 4-5.

**Table 1.**
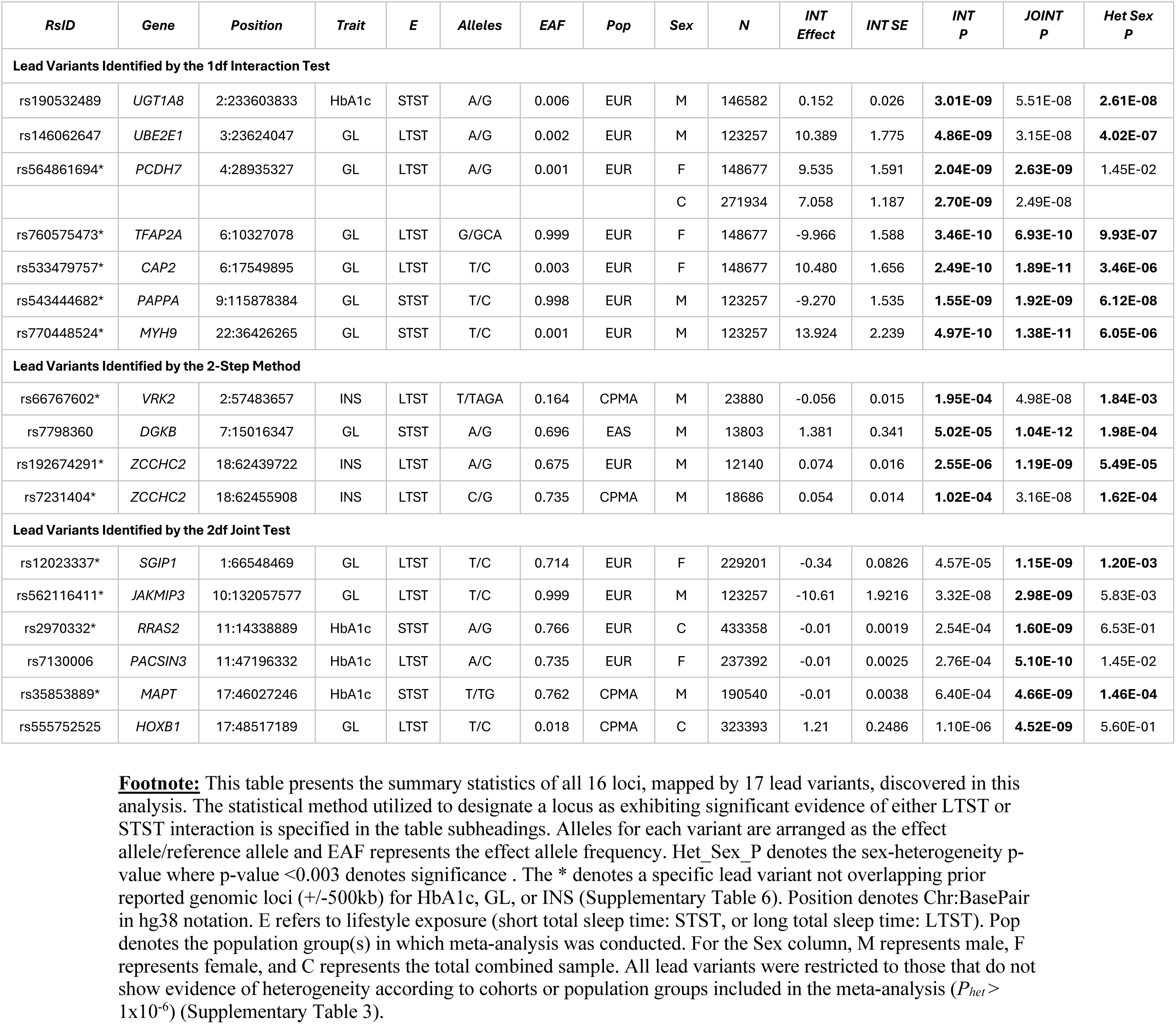
Gene-sleep duration interaction loci for glycemic traits.

Among the 17 lead variants, nine were common variants (MAF>1%) identified in cross-population meta-analysis (CPMA; 4), EAS meta-analysis (1), or EUR meta-analysis (4). The remaining eight relatively rare variants (MAF between 0.1% and 1%) were identified only in the UK Biobank. Majority of the 17 lead variants were intergenic (12) with five variants overlapping introns (4) or the 3’UTR (1) of protein-coding genes. Thirteen of the 17 lead variants (corresponding to 12 independent loci) showed evidence of sex-dependent effects (*P_sex_het_*< 3×10^-3^).

### Cross-Population Meta-Analysis (CPMA)

Four of the common lead variants (MAF>1%) were identified from CPMA. Combined sex CPMA for GL revealed rs555752525 (proximal to *HOXB1*; *P_M1_G×E_*=1.1×10^-6^) to interact with LTST by the 2df joint test. Male CPMA for INS revealed rs66767602 (proximal to *VRK2*; *P_M1_G×E_*=1.95×10^-4^) and rs7231404 (proximal to *ZCCHC2*; *P_M1_G×E_*=1.02×10^-4^) to interact with LTST by the two-step method. Male CPMA for HbA1c revealed rs35853889 (3’UTR variant of *MAPT*; *P_M1_G×E_*=6.40×10^-4^) to interact with STST by the 2df joint test.

### Population Specific Meta-Analysis

EAS or EUR population specific meta-analyses identified the remaining five common (MAF>1%) lead variants. Male EAS meta-analysis for GL identified rs7798360 (proximal to *DGKB*; *P_M1_G×E_*=5.02×10^-5^) to interact with STST by the two-step method. Male EUR meta-analysis for INS identified rs192674291 (proximal to *ZCCHC2*; *P_M1_G×E_*=2.55×10^-6^) to interact with LTST by the two-step method. This locus overlapped (+/- 500 kilobases) the aforementioned male CPMA rs7231404 INS-LTST locus. Combined sex EUR meta-analysis for HbA1c revealed rs2970332 (intronic variant of *RRAS2*; *P_M1_G×E_*=2.54×10^-4^) to interact with STST by the 2df joint test. The 2df joint test in female EUR meta-analysis for GL revealed rs12023337 (intronic variant of *SGIP1*; *P_M1_G×E_*=4.57×10^-5^) to interact with LTST, and for HbA1c, revealed rs7130006 (proximal to *PACSIN3*; *P_M1_G×E_*=2.76×10^-4^) to interact with LTST.

All eight relatively rare lead variants (MAF <= 1%) were identified in a single large EUR study, the UK Biobank. For the GL meta-analysis, the 1df interaction test identified four variants interacting with LTST: rs146062647 (proximal to *UBE2E1*; *P_M1_G×E_*=4.86×10^-9^), rs533479757 (intronic for *CAP2*; *P_M1_G×E_*=2.49×10^-10^), rs564861694 (proximal to *PCDH7*; *P_M1_G×E_*= 2.70×10^-9^; *P_M1_G×E_females_*= 2.04×10^-9^), rs760575473 (proximal to *TFAP2A*; *P_M1_G×E_*=3.46×10^-10^), and two variants interacting with STST: rs543444682 (proximal to *PAPPA*; *P_M1_G×E_*=1.55×10^-9^) and rs770448524 (proximal to *MYH9*; *P_M1_G×E_*=4.97×10^-10^). For HbA1c meta-analysis, the 1df interaction test revealed rs190532489 to interact with STST (proximal to *UGT1A8*; *P_M1_G×E_*=3.01×10^-9^). The 2df joint test identified rs562116411 (intronic variant of *JAKMIP3*; *P_M1_G×E_*=3.32×10^-8^) to interact with LTST for GL. rs190532489, rs146062647, rs770448524, rs562116411, and rs543444682 were identified in males, and rs533479757 and rs760575473 in females. rs564861694 was identified in both combined sex and females.

### Overlap with Glycemic Loci

Of the 16 prioritized interaction loci (mapped by 17 lead variants), 11 loci were revealed to be novel for glycemic traits. Among the other five, three loci overlapped (+/-500kb) previously reported HbA1c or fasting GL GWAS loci: rs146062647 (*UBE2E1*) for HbA1c; and rs7798360 (*DGKB*) and rs555752525 (*HOXB1*) for fasting GL (Supplementary Table 3, Supplementary Table 6) (15). rs190532489 (*UGT1A8*) and rs7130006 (*PACSIN3*) overlapped prior reported GWAS loci for other glycemic traits, including measures of random glucose, insulin response, or fasting proinsulin (Supplementary Table 3, Supplementary Table 6) (15).

### Variant Annotation

The 17 lead variants were mapped to a total of 16 primary protein-coding genes based on either direct positional overlap (e.g., intron, 3’ UTR), or nearest distance to transcription start site.

Nine variants exhibited high regulatory potential by RegulomeDB (categories 1f, 2b; Supplementary Table 3, Supplementary Table 7) (16). In total seven lead variants showed expression quantitative trait loci (eQTL), splicing QTL (sQTL) or protein QTL (pQTL) evidence (*P*<0.05) in adipose, pancreatic, and/or brain tissues (Supplementary Table 3). Specifically, rs2970332 and rs7130006 exhibited pQTL associations in prefrontal cortex tissue; and rs192674291, rs7231404, rs12023337, rs7130006, rs35853889 and rs555752525 eQTL associations in pancreatic tissue. In addition, rs192674291, rs7231404, rs12023337, rs35853889, and rs7130006 showed sQTL or eQTL associations in brain tissue; and rs192674291, rs7231404, rs12023337, rs7130006, rs35853889, and rs555752525 exhibited sQTL or eQTL associations in adipose tissue.

Three of the 17 lead variants were found to exhibit strong (*P*<5×10^-8^) associations with glycemic and type 2 diabetes-related traits in prior GWAS curated by the Type 2 Diabetes Knowledge Portal (T2DKP) (Supplementary Table 7) (15). Specifically, rs7798360 is associated with type 2 diabetes, diabetic retinopathy, and type 2 diabetes with neurological manifestations. Variants rs7798360, rs2970332, and rs35853889 are associated with glucose measures, insulin response, or HbA1c (15). T2DKP, Common Metabolic Diseases Knowledge Portal (CMDKP) and GWAS Atlas additionally revealed the following metabolic or cognitive-related lead variant associations: rs190532489 (bilirubin), rs7231404 and rs192674291 (calcium, alkaline phosphatase, albumin), rs2970332 (vitamin D), and rs7130006 (alkaline phosphatase, albumin, feeling fed-up, depressed affect, miserableness) (17; 18).

### Gene Annotation

The 16 primary genes (mapped to the 17 lead variants) were annotated for their reported significant associations (*P*<5×10^-8^) with complex traits or disorders (Supplementary Table 8). Most relevant to glycemic and sleep health, *VRK2*, *UGT1A8*, *UBE2E1*, *DGKB*, *PAPPA*, *MYH9*, *SGIP1*, *JAKMIP3*, and *MAPT* are associated with glucose, HbA1c, insulin, metabolic syndrome, type 2 diabetes, diabetes complications (diabetes with ketoacidosis, polyneuropathy), or diabetes medication use. *VRK2*, *UGT1A8*, *DGKB*, *ZCCHC2*, *SGIP1*, *MAPT*, and *HOXB1* are associated with sleep duration, daytime nap, chronotype, insomnia, or ease of waking. Extending to animal model evidence, gene knockouts in mice for *CAP2*, *SGIP*1, *MYH9*, and *JAKMIP3* alter triglyceride levels, aspartate transaminase, or lipase levels (Supplementary Table 8) (19). Knockouts of *RRAS2* and *ZCCHC2* in mice are reported to induce changes in body mass, body weight, or body fat amount (19). Lastly, humans genetic variation catalogued in OMIM (https://omim.org/) for neurological, hematological, or cardiovascular mendelian disorders implicate *CAP2* for dilated cardiomyopathy, *MYH9* for macrothrombocytopenia and granulocyte inclusions, and *MAPT* for multiple neurological disorders including Parkinson’s disease, and frontotemporal dementia (Supplementary Table 8).

### Enrichment Analysis

The expanded set of genes mapped to each lead variant locus according to distance, quantitative trait loci, or chromatin interaction evidence (Supplementary Table 3), was investigated by STRING and FUMA GENE2FUNC gene set-based enrichment analyses (FDR<0.05) (Supplementary Figure 6, Supplementary Tables 9-10) (20; 21). GL loci interacting with STST was enriched in dietary heme iron intake interaction for Type 2 diabetes, residual cognition, and total intracranial volume. GL loci interacting with LTST was enriched in similar traits of metabolism (regulation of food intake, BMI) and neurological health (epilepsy, high intelligence). HbA1c loci interacting with LTST were enriched in short sleep duration and metabolic traits (e.g., HDL cholesterol, BMI), psychiatric health (e.g., mood instability, neuroticism), and Alzheimer’s disease. HbA1c loci with LTST interaction were enriched in neurological phenotypes (e.g., Alzheimer’s disease, white matter integrity), psychiatric symptoms (e.g., neuroticism, irritability), metabolic traits (e.g., serum urea, bilirubin, BMI), and vitamin levels (vitamin D). Differing from the GL-LTST and HbA1c-LTST enrichment results, INS-LTST loci highlighted serine-type endopeptidase inhibitor activity and epithelial cell adhesion molecule levels.

Analyzing genome-wide 1df G×E summary statistics by Multi-marker Analysis of GenoMic Annotation (MAGMA), identified *SLFN12*, *C19orf80*, and *RAB27A* to be significantly enriched in gene-based analysis (Supplementary Table 11) (22). MAGMA gene-set enrichment analysis additionally revealed enrichment in lipid metabolism (ceramide 1-phosphate transfer activity, adipogenesis), and cellular dynamics (DNA Double strand break repair, SNARE complex assembly, oxidation reactions) (Supplementary Table 11).

### Therapeutic Connections

For insights into clinical application, primary genes were investigated for whether they are targets of, or implicated in the efficacy of existing drug or therapeutics (Supplementary Table 8). This identified *UGT1A8* to be implicated in the glucuronidation of several therapeutics prescribed for bipolar disorders or seizures (lamotrigine), pain reduction (naproxen, diflunisal, morphine, acetaminophen), opioid overdose (nalmefene), type 2 diabetes, chronic kidney disease, and heart failure (empagliflozin), and neurological disease (edaravone). *PAPPA* is denoted to be targeted by phenethyl isothiocyanate and *UGT1A8* inhibited by curcumin and cannabinoids. Genetic variation mapped to *CAP2* is associated with selective serotonin reuptake inhibitor response and variation mapped to *DGKB* associated with fluctuations in weight from antipsychotics.

Extending analysis to the broader set of genes mapped to the 16 interaction loci (Supplementary Table 3), revealed additionally five genes to be established targets of FDA-approved drugs (Supplementary Tables 12-13). *JAK1* (*rs12023337* locus) is a target of several small molecule inhibitors (e.g., tofacitinib), approved to treat patients with autoimmune diseases such as rheumatoid arthritis. *F2* (*rs7130006* locus) is a target of anticoagulants (e.g., argatroban), used to treat heparin-induced thrombocytopenia, acute coronary syndrome and venous thromboembolism. *LEPR* (*rs12023337* locus) is a target of adjunct diet replacement therapy (metraleptin) to treat patients with lipodystrophy. Two well-established targets of phosphodiesterase inhibitors — *PDE3B* (rs2970332 locus) and *PDE4B* (rs12023337 locus) — are used to treat congestive heart failure, pulmonary diseases, and smooth muscle contractility (e.g., inamrinone, iloprost, papaverine).

## Discussion

### Overview

We conducted the first genome-wide gene-by-short and long sleep duration interaction study for three glycemic traits in up to 489,309 individuals without diabetes mellitus. In total 16 genomic loci exhibit interactions with short or long sleep duration, including 11 loci novel to glycemic traits (*VRK2, PCDH7, TFAP2A, CAP2, PAPPA, ZCCHC2, MYH9, SG1P1, JAKMIP3, RRAS2, MAPT)*. Multiple loci (15) were specific to sex and/or particular (EAS, EUR) population groups, suggesting heterogeneity is present in variant-sleep duration interaction effects.

### Neurological Connections

Similar to prior gene by sleep interaction studies for blood pressure and blood lipids, many of the mapped genes are linked to neurological health (Supplementary Tables 7-8), highlighting the role of the central nervous system — the fundamental regulator of sleep — in influencing insulin and glucose levels through neuronal and neuroendocrine pathways that control eating, energy expenditure and metabolism (9; 23). In this perspective, primary genes identified by lead variants in this study — *PCDH7, TFAP2A,* and *MAPT* — appear to be notable novel targets for type 2 diabetes, exhibiting dual roles in both sleep health, and glycemic regulation.

Firstly, *PCDH7* encodes a protocadherin that binds to GluN1, a subunit of the NMDA receptor (24). GluN1 is critical for healthy sleep, with loss of GluN1 in the lateral preoptic hypothalamus able to induce insomnia and fragmented NREM sleep in mouse models (25). NMDA receptors are a therapeutic target for eating disorders, and their pharmaceutical targeting may carry further benefits for glycemic control – illustrated by an investigational dual-action drug (GLP-1–MK-801) with NMDA receptor antagonism and GLP-1 receptor agonism shown to alleviate both hyperglycemia and obesity (26). Additionally, given *PCDH7*’s reported associations with short sleep duration, neuron dendrite morphology, and the regulation of synaptic NMDA receptor currents, future work investigating how sleep duration affects *PCDH7* gene expression, and in turn downstream effects may be valuable (24; 27). Secondly, *TFAP2A* has been shown to regulate sleep quality and circadian period in rodent models, with its reduction in expression resulting in stronger delta and theta EEG power during sleep (28). This is important because delta is the dominant EEG component of slow-wave sleep — a sleep stage whose suppression is correlated with loss of insulin sensitivity, and is diminished in obesity (29). Lastly, *MAPT*, which encodes the protein tau, is a well-established target of several neurodegenerative diseases. *MAPT* is also expressed in pancreas islets, where it assists in insulin granule exocytosis, with studies having demonstrated its expression to influence both insulin transcription and secretion rates (30). Noting that *MAPT* genetic variation has been attributed to multiple type 2 diabetes complications (neuropathy, retinopathy, renal), and to multiple sleep traits (ease of waking in the morning, daytime napping, narcolepsy, sleep duration, and snoring), investigating the role of sleep health on *MAPT* expression and type 2 diabetes pathogenesis warrants further investigation (Supplementary Table 8).

### Novel Pathway Connections to Type 2 Diabetes

Beyond dual sleep-glycemic connections, important cellular mechanisms in adipocytes specifically pertaining to type 2 diabetes pathogenesis were highlighted by two primary genes identified in this study: *PCDH7,* and *PACSIN3.* In adipocytes, *PACSIN3* modulates glucose uptake by regulating the trafficking of the GLUT1 receptor (31). Posttranslational modification may be a key to its function, with phosphorylation of its specific serine residues (268 and 354) proposed to regulate *PACSIN3* activity on GLUT1, in turn influencing adipose glucose transport (31). Extending to cellular interactions, under exposure to diabetes serum, pericytes – critical for vascular integrity – exhibit dampened *PCDH7* expression in adipocytes (32). Compromise of key cell adhesion proteins like *PCHD7* can substantially weaken pericyte-endothelial cell interactions, disrupt blood vessel integrity, and promote apoptosis of pericytes (32). Pericyte apoptosis may precede diabetes onset, as mice models with depletion of pancreatic pericytes show impairment of beta cell glucose-stimulated insulin secretion, and loss of beta cell maturity (32; 33).

### Molecular Mechanisms Specific to Short or Long Sleep Duration

The nonoverlap between loci interacting with short or long sleep indicates distinct mechanisms influencing type 2 diabetes. In this context, it is noteworthy that select LTST (*PACSIN3*) and STST interaction (*DGKB)* loci highlight two distinct molecular pathways by which long sleep or short sleep may act upon glycemic traits: copper and diacylglycerol metabolism.

Hyperglycemia can increase cellular buildup of both advanced glycosylation end products (AGEs) and copper, eventually leading to cuproptosis, a form of triggered cell death implicated in diabetic cardiomyopathy (34). Dysregulated copper homeostasis, through its effects on oxidative stress, inflammation, and energy metabolism, has been implicated in the development of type 2 diabetes (35). Sleep may play a role in copper dynamics, as sleep fragmentation has been found to induce copper overload in cardiomyocytes and long sleep duration positively correlated with serum copper (36; 37). Given this context, *SPI1* – a transcription factor mapped to the *PACSIN3* locus (Supplementary Table 3) – regulates SLC31A1, the copper importer upregulated by AGEs (38). Considering *PACSIN3*’s aforementioned ability to regulate the GLUT1 receptor, it may be important for future studies to investigate the role of long sleep duration in influencing copper dynamics, and *PACSIN3* protein interactions, for glycemic regulation.

For short sleep duration, a direct link to cellular diacylglycerol (DAG) metabolism was implicated by *DGKB*, encoding a kinase that regulates intracellular DAG levels. DAG is reduced in serum after acute sleep restriction, and DAG accumulation in the liver is linked to insulin resistance (39; 40). Furthermore, the serum of mice with prediabetes show the presence of DAGs, with persistently higher levels in diabetes (41). Melatonin – the pituitary hormone critical for regulating circadian rhythm – has also been shown to modulate levels of diacylglycerol in human blood lymphocytes (42). Especially given that *DGKB* has been associated with several glycemic traits, type 2 diabetes, and insomnia in prior GWAS (Supplementary Tables 7-8), it may be important to analyze how sleep and circadian rhythm influence DAG fluctuations, and in turn insulin resistance.

### Opportunities for Therapeutic Design or Repurposing in Type 2 Diabetes

Several of the identified genes point to potentially promising therapeutic design or drug repurposing strategies for type 2 diabetes or its complications. For instance, argatobran (targets *F2*) has been demonstrated in rodent models to potentially treat diabetic cardiomyopathy, and metraleptin (targets *LEPR*) shown to improve HbA1c levels and insulin response in patients with lipodystrophy (43; 44). Second, *VRK2* has been shown to disrupt the interaction between JIP1 scaffold protein and JNK by binding to the C-terminal region of JIP1 (45). The disruption of JIP1-JNK interaction is a proposed treatment strategy to restore insulin sensitivity (46). Third, a monoclonal antibody targeting *SERPINB13* (mapped to *ZCCHC2* locus) has been shown to promote proliferation of insulin-producing pancreatic islet cells and reduce inflammation (47). Fourth, phenethyl isothiocyanate (found in cruciferous vegetables) is able to suppress appetite and enhance leptin signaling (48). *PAPPA* is targeted by this phytochemical as well as resveratrol, a polyphenol with well-established cardioprotective and hypoglycemic properties (49; 50).

### Strengths and Limitations

This study has several notable strengths. It represents the first large-scale gene-by-sleep interaction analysis for glycemic traits, leveraging data from numerous international biobanks and cohorts that include diverse population groups. We undertook extensive efforts to address heterogeneity across data sources through rigorous harmonization protocols to ensure the robustness and reproducibility of our findings. Additionally, we conducted sex- and population-stratified analyses to further explore effect heterogeneity. Lastly, we applied multiple bioinformatics approaches to deepen our understanding of the biological mechanisms involved and to identify promising therapeutic targets for glycemic control.

This study also has several limitations. First, the exclusive use of self-reported sleep data may be subject to recall bias and misclassification. Notably, long sleep may be partly influenced by spending longer time in bed rather than sleeping longer. Moreover, sleep health is a multidimensional construct that extends beyond duration. Future studies employing objective measures such as actigraphy to capture these broader dimensions are needed to better understand the specific aspects of sleep and circadian rhythms that moderate risk for metabolic dysfunction. Second, despite our inclusion of international cohorts and biobanks representing a broad range of populations, the sample remains predominantly of European ancestry. Future replication, especially in non-European groups, is warranted. Third, to reduce analytic burden across multiple sites, we applied a minimally adjusted model including age, sex, and population structure. As a result, there may be residual confounding from unmeasured behavioral and clinical factors (e.g. comorbid disorders such as sleep apnea or anemia). Finally, this study focused solely on cross-sectional interactions with glycemic traits, which does not imply causality. Longitudinal G×E analyses using time-to-event data on type 2 diabetes incidence will be critical for informing personalized risk mitigation strategies.

### Conclusion

In summary, we identified 16 genomic loci across three glycemic traits that exhibit significant interactions with either short or long total sleep duration. Our findings suggest that short and long sleep durations influence glycemic regulation through neurological regulation and other distinct biological pathways influencing diacylglycerol and copper levels, offering novel insights that may inform precision strategies for diabetes prevention and management.

## Data Availability

Genome-wide summary statistics of the cross-population gene-sleep duration interaction analyses will be made available in the GWAS Catalog. Source code for primary software used to conduct GxE meta-analysis is publicly available at the following repositories: GEM (https://github.com/large-scale-gxe-methods/GEM), MMAP (https://mmap.github.io/), LinGxEScanR (https://github.com/USCbiostats/LinGxEScanR), FUMA (https://fuma.ctglab.nl/), METAL (https://github.com/statgen/METAL).

## Acknowledgements

Study-specific acknowledgements are included in the Supplementary Note.

## Article Information

### Author Contributions

H.W. is the guarantor of this work and, as such, had full access to all the data in the study and takes responsibility for the integrity of the data and the accuracy of the data analysis. H.W. and P.N. conducted core project data consolidation, data cleaning, data analysis, bioinformatics analysis, and result interpretation. H.W., P.N., C.L.M., A.R.B., R.N., K.E.W., D.C.R., A.K.M., H.C., L.d.l.F., S.R., and J.B.M. formed the writing group that closely worked on interpretation of results, and drafting the manuscript. L.d.l.F., A.D., C.G., and D.C.R. were involved in centralized study coordination. H.W., P.N., K.E.W., D.C.R., A.K.M., and J.B.M. worked on consolidation of genomic loci for novelty annotations. A.T.K., J.L.M., J.R.O., T.W.W., H.A., J.B.M., X.Z., H.C., J.G., A.K.M., C.L.M., P.B.M., and P.A.P. assisted in design of computational workflows. All other co-authors participated in cohort-level phenotype and genotype data acquisition, cohort-level data analysis, and final interpretation of results. All authors approved the final version of the paper that was submitted to the journal.

