## Supplementary Information for "Gene-by-Sleep Duration Interaction for Glycemic Traits in over 480,000 Individuals"

*Wang H, et al.*

### Supplementary Methods

#### Study Design

Population groups as a label for underlying ethnicity, ancestry and/or geographical origin were defined African (AFR), Admixed American (AMR), East Asian (EAS), European (EUR), Hispanic/Latinos (HIS), Middle Eastern (MID), and South Asian (SAS). These groups were carefully considered by individual study teams to encompass both ethnic identity and genetic similarity for pre-meta-analysis quality control and post-meta-analysis linkage disequilibrium-based variant pruning. For most of the cohorts, samples were mapped to AFR, EAS, EUR, HIS, or SAS groups, based on self-reported identity or individuals closely clustering with one of the 1000 Genome Phase 3 super population (Supplementary Table 1). UK Biobank (UKB) participants were mapped to AFR, AMR (sharing ancestry with many HIS groups), EAS, EUR, MID (sharing ancestry background with EUR), and SAS populations by the Pan-UKB project, based on genetic similarity to population groups from two reference datasets: the 1000 Genomes Project and the Human Genome Diversity Project (HGDP) [https://pan-dev.ukbb.broadinstitute.org/docs/study-design/]. For quality control and linkage disequilibrium panel comparisons, MID was mapped to 1000G EUR based on close clustering via principal component analysis (PMID: 40968291) and HIS was mapped to 1000G AMR due to this reference panel reflecting Colombian, Peruvian, Mexican, and Puerto Rican individuals.

A standardized analysis plan was circulated to 30 participating studies (two biobanks; 28 cohorts), resulting in a total of 52 population group-specific study summary statistics submitted to the central project team for quality control, meta-analysis, variant prioritization, and bioinformatics interpretation. Each cohort conducted regression analyses according to models M1 and M2 defined below, both within specific population groups (AMR, AFR, EAS, EUR, HIS, MID, SAS), and sex (males, females, combined sex), as applicable to the data. Y denotes the glycemic trait, E denotes a lifestyle exposure (STST or LTST), G is a SNP, and *C_1_* includes age, age^2^, sex, age×E, age^2^×E, sex×E, genomic principal components (PCs), and cohort-specific covariates (e.g. study center). C_2_ denotes age, age^2^, sex, genomic PCs, and cohort-specific covariates. For sex-specific analysis, covariate vectors C_1_ and C_2_ did not include sex.

Cohort-specific covariates and descriptive summary statistics are provided in Supplementary Table 1. Each team used the following analysis software: MMAP (<https://mmap.github.io/>), LinGxEScanR (<https://github.com/USCbiostats/LinGxEScanR/tree/CHARGE>) and/or GEM (<https://github.com/large-scale-gxe-methods/GEM>). Longitudinal studies were instructed to select a single visit based on maximized sample size. Studies with known sample relatedness or suspected cryptic relatedness were asked to use residuals adjusting for either a kinship matrix or genetic covariance matrix as the quantitative outcome for analysis. Genotype data was restricted to autosomal chromosomes 1-22, imputation quality >=0.3 and minor allele frequency (MAF) >=0.1%.

Model 1 (Primary GxE Model of Interest)

M1: Y = B_M1_0_ + B_M1_G_G + B_E_E + B_GxE_GxE + B_M1_C_ C_1_

Model 2 (Marginal Genetic Effect Model for Comparison)

M2: Y = B_M2_0_ + B_M2_G_G + B_M2_C_ C_2_

#### Phenotypes

The three glycemic traits (Y) analyzed were HbA1c (%), Glucose (GL; mg/dL), and Insulin (INS; µU/mL).

Glycemic traits are typically collected in cohort studies with standard research protocols which usually require study participants to observe fasting for ≥ 8 hours before visiting the clinics for data collection. Whereas HbA1c can be measured in total samples (regardless of fasting status), GL and INS were considered when measured in fasting samples only. Large biobanks usually rely on Electronic Health Records (EHR), without the elaborate protocols typical of cohort studies. In particular, most of the EHR data may not comply with fasting requirements. Furthermore, INS is not routinely checked in the clinical setting (ie, EHR-based biobanks) for the screening, diagnosis, or management of diabetes. Therefore, when insulin levels are measured outside of the research environment (i.e. for suspicion of disease or disease management), they typically represent dramatic outliers (e.g. suspicion of endocrine cancers or endocrine failure). Therefore, we designed separate harmonization plans for cohort studies and biobanks.

In cohort studies, we analyzed HbA1c in Fasted + Not-Fasted combined samples, and GL and INS in reported fasting samples only. In biobanks, INS was not analyzed, HbA1c was analyzed in Fasted + Non-Fasted combined samples, and GL was analyzed in fasted (GL-F) and not-fasted (GL-NF) samples separately. GL and INS collected using other units were converted into a standard unit first (1 mmol/L = 18 mg/dL and 1 pmol/L = 0.16 μU/mL). Fasting INS in cohort studies was natural logarithm transformed and adjusted for BMI. Prior work has demonstrated that adjusting for BMI for INS can improve genomic loci discovery [PMID: 34059833]. Trait outliers (>6 standard deviations above or below the mean) were winsorized. Participating samples were restricted by age (>=18 years), total self-reported sleep time (>=3 and <=14 hours), hospitalization status (not marked as undergoing inpatient care), and diabetes status (no present diabetes according to individual study definitions, and/or HbA1c >=6.5% or GL >=126 mg/dL in fasting cohorts and GL_F, and HbA1c >=6.5% or GL >=200 mg/dL in GL-NF).

Two binary (1/0) environment (E) terms were tested for interaction with genetic variants (G): short total sleep time (STST) and long total sleep time (LTST). 20^th^ and 80^th^ percentiles from total sleep time regressed on age, sex, and age x sex were denoted as STST and LTST respectively (STST=1 if <=20^th^ percentile; LTST=1 if >=80^th^ percentile).

Summary statistics of variants (sample size, genomic position, allele descriptors, imputation quality) and statistical test results (P-values, model-based standard errors, effect size, covariance), were deposited centrally to the meta-analysis team for the following: 1 degree of freedom (1df) marginal (M2_G) genetic effect (H_0_: B_M2_G_ = 0), 1df main genetic (M1_G) effect (H_0_: B_M1_G_ = 0), 1df GxE interaction effect (H_0_: B_M1_GxE_ = 0), and 2df joint G,GxE effect (H_0_: B_M1_G_ = 0;B_M1_GxE_ = 0).

#### Genotype and Quality Control

Prior to meta-analysis EasyQC2 software (v1.1.2) was first used to clean the summary statistics files at the study-level (https://homepages.uni-regensburg.de/~wit59712/easyqc2/charge2qc_240424.ecf). Rows with missing values, monomorphic variants, invalid out-of-range statistics, and extreme effect sizes (>1000) were filtered out. Results that did not reflect minor allele count * imputation quality >=20 in the exposed (E=1), unexposed (E=0), and entire sample were excluded. All variants were converted to GRCh38 coordinates. Quantile-Quantile plots of p-values were assessed for striking inflation or deflation warranting errors to fix and rerun analysis at the cohort level. Trans-Omics for Precision Medicine (TOPMed)-imputed 1000 Genomes Phase 3 v5 reference panels were used to assess reported allele frequencies for any alarming discrepancies. The output cleaned files were then assessed at the group-level (binned according to population groups), assessing for problematic trait transformations (e.g. outcome centering) or striking outlier studies (<https://homepages.uni-regensburg.de/~wit59712/easyqc2/charge2premetaqc_231214.ecf>).

#### Inverse-Variance Weighted Meta-Analysis

Following quality control, for meta-analysis cohorts were defined to be stratified according to the seven population groups (AFR, AMR, EAS, EUR, HIS, MID, SAS) and three sex groups (male sex, female sex, combined sex). Any given GL, INS, or HbA1c meta-analysis was required to have to at least 20,000 samples or 3 contributing studies to warrant downstream variant prioritization and results reported. METAL software (release 2010-02-08) with inverse variance weights via SCHEME STDERR for 1df tests (β_M1_G_, β_M2_G_, β_M1_GxE_ ) and SCHEME INTERACTION for the 2df test (β_M1_G_ ; β_M1_GxE_ ) was utilized to meta-analyze summary statistics, within each population group. (PMID: 21181894). Following this, for cross-population meta-analysis (CPMA) the output results from these population group-specific meta-analyses were then meta-analyzed. For all METAL runs, genomic correction was enforced to correct for inflation (GENOMICCONTROL ON) and heterogeneity tests conducted (ANALYZE HETEROGENEITY).

#### Significant Interactions

EasyStrata2 software (v1.2.7) was used to specify independent genomic loci from significant variants. All significant variants were restricted to those that do not show evidence of heterogeneity (P_het_ > 10e-6 ). The genomic region encompassing the major histocompatibility complex region (chr6: 27500000-3450000) was excluded. Variants harboring evidence of LTST or STST interaction were determined by three methods: (A) 2-step method, (B) 1df GxE test, and (C) 2df G,GxE joint test. Method (A) is as follows. First variants with M2_G<1e-5 are used to calculate effective number of tests by principal component analysis, denoted as *m_G_*. Following this, variants with Bonferroni-adjusted P_GxE_2step_ < 0.05 (corrected for multiple tests according to 2**m_G_*) are denoted significant. Method (B) denotes variants that pass P_GxE_ < 5e-08. Method (C) denotes variants that pass P_G,GxE_ < 5e-08. Variants identified by (A) or (B), and (C) were each pruned to independent lead variants that tag neighboring variants defined by LD threshold r^2^<0.1 within 500 kilobase distance windows, using TOPMed-imputed 1000 Genomes reference panels. For any variants missing in the LD reference panel used, if such a variant was the top significant signal in a given 500 kilobase region, it was retained as the lead variant.

From this set of significant loci, variants were then filtered to a final set of lead variants that pass prioritization criteria, V*. Specifically, Method (A) lead variants were restricted to those that pass FDR__GxE_2step_ <0.05. Method (B) lead variants were restricted to those that pass P_GxE_<5e-09 and FDR_GxE_<0.05. Method (C) lead variants were restricted to those with P_G,GxE_ < 5e-09 , exhibit insignificant marginal genetic effect (P_M2_G_ >=5e-08), and stronger interaction signal relative to the main genetic effect (P_GxE_ < P_M1_G_). Lastly, the Type 2 Diabetes Knowledge Portal resource was used to retrieve a set of carefully curated genomic loci for glucose, HbA1c, and insulin traits (see Supplementary Table 4). Using this curated list, lead variants were annotated for overlap (+/-500kb) with prior reported loci for fasting glucose, HbA1c, fasting insulin, and other glycemic traits.

#### Heterogeneous Effect According to Sex

To assess sex-specific heterogeneity, two-sample Z-tests assuming independence when comparing males to females, were conducted for each of the V* prioritized variants, with significance determined as P<0.05/V*.

#### Gene Mapping

Each lead variant from V* was mapped to a primary gene (G^**^), and an extended set of mapped genes (G^~^). For downstream clinical interpretation and druggability analysis, all genes were restricted to protein-coding. Primary genes were determined by direct gene overlap (e.g. exonic) or nearest distance to canonical transcription start site (TSS) using Ensemble Release 113 and APPRIS annotations. Extended sets of mapped genes were those identified by a multi-evidence strategy (1) FUMA SNP2GENE (v1.6.2), (2) Open Targets Genetics, (3) QTLbase and (4) eQTLGen. For (1) FUMA SNP2GENE mapping, the following three settings were used. Firstly, for positional mapping: protein-coding genes were mapped if <=10kb to the lead variant, based on ANNOVAR annotation. Secondly, expression quantitative trait loci (eQTL) associations implicated genes whose expression is associated (FDR<0.05) with either the lead variant, or variants in LD with the lead variant (R2>=0.6 based on 1000G Phase 3 reference panels). Lastly, via chromatin interaction mapping, genes were mapped if their promoter regions (250 bp upstream of TSS to 500 bp downstream of TSS] were associated (FDR<1e-6) with the lead variant or variants in LD with the lead variant (same LD criteria as above for eQTL). For (2) Open Targets Genetics mapping, genes identified by top Variant-to-Gene (V2G) score, and those mapped by eQTL, protein QTL, and/or chromatin interactions were noted. For (3) QTLbase mapping, genes implicated by significant (p<0.05) pQTL associations within +/- 10Mb of lead variants were noted. For (4) cis-eQTLs within +/- 1Mb were retrieved.

#### Variant Annotations

All lead variants from V* were annotated according to most severe consequence reported by Ensembl Variant Effect Predictor (VEP), CADD score (gnomAD v4.1.0), RegulomeDB (v2.2) (regulatory score; high transcription activity chromHMM marks; transcription factor binding site/motif overlap; GTEXv8 eQTL), and significant trait associations (p<5e-08) reported by multiple databases (Open Targets Genetics; PheWeb: https://pheweb.org/UKB-TOPMed/; Common Metabolic Disorders Knowledge Portal; Sleep Disorders Knowledge Portal; PheGenI; LDtrait: [R2>=0.6, 500kb window, ALL populations, GRCh37 Genome Build coverage]; GWAS catalog; GWAS Atlas).

#### Primary Gene Annotations

G** mapped by V* were each annotated according to significant (p<5e-08) trait associations (Open Targets Genetics: L2G score >=0.8; PheWeb; GWAS Atlas; GWAS Catalog; PheGenI; Common Metabolic Disorders Knowledge Portal: HUGE score >=30; Sleep Disorders Knowledge Portal: HUGE score >=30), druggability evidence (DrugBank, PharmGKB, Drug-Gene Interaction Database (DGIdb v5.0.8): interaction score >=1), animal model gene knockout functional annotations (International Mouse Phenotyping Consortium (IMPC)), and human Mendelian disorders (https://www.omim.org/).

#### Gene-Based and Gene-Set Enrichment Analysis

MAGMA tissue-enrichment, gene set enrichment and gene-based analysis was run using the FUMA SNP2GENE platform, with 1df GxE meta-analysis results provided as input. FUMA GENE2FUNC was used to query G~ in sex-exposure groups M* (e.g. LTST-Females) for significant (Bonferroni adjusted p-value< 0.05) enriched pathways, traits, or ontology terms from a protein coding gene background set. STRING (v12.0) was used to build protein-protein interaction (PPI) networks from M* and identify significantly enriched (FDR<0.05) pathways, ontology terms, and traits.

#### Druggability Analysis

We first used the Drug-Gene Interaction database (DGIdb; v4.2.0) to query high or medium priority sleep-glycemic trait interacting genes to determine the potentially druggability of the candidate gene targets. We annotated genes for implicated pathways and functions using the Kyoto Encyclopedia of Genes and Genomes (KEGG) database. We annotated the druggability target categories and queried all interacting drugs reported in 43 databases (BaderLabGenes, CarisMolecularIntelligence, dGene, FoundationOneGenes, GO, HingoraniCasas, HopkinsGroom, HumanProteinAtlas, IDG, MskImpact, Oncomine, Pharos, RussLampel, Tempus, CGI, CIViC, COSMIC, CancerCommons, ChemblDrugs, ChemblInteractions, ClearityFoundationBiomarkers, ClearityFoundationClinicalTrial, DTC, DoCM, DrugBank, Ensembl, Entrez, FDA, GuideToPharmacology, JACX-CKB, MyCancerGenome, MyCancerGenomeClinicalTrial, NCI, OncoKB, PharmGKB, TALC, TEND, TTD, TdgClinicalTrial, Wikidata). We queried protein targets for available active ligands in ChEMBL. We queried gene targets in the druggable genome using the most recent druggable genome list established from the NIH Illuminating the Druggable Genome Project (https://github.com/druggablegenome/IDGTargets) available through the Pharos web platform. We also queried FDA-approved drugs, late-stage clinical trials and disease indications in the DrugBank, ChEMBL, ClinicalTrials.gov databases and provided results for the top MESH and DrugBank indications and clinical trials.

### Supplementary Notes

#### Study Descriptions

**Age Gene/Environment Susceptibility Reykjavik Study (AGES):** The AGES Reykjavik study originally comprised a random sample of 30,795 men and women born in 1907-1935 and living in Reykjavik in 1967. A total of 19,381 people attended, resulting in a 71% recruitment rate. The study sample was divided into six groups by birth year and birth date within month. One group was designated for longitudinal follow up and was examined in all stages; another was designated as a control group and was not included in examinations until 1991. Other groups were invited to participate in specific stages of the study. Between 2002 and 2006, the AGES Reykjavik study re-examined 5,764 survivors of the original cohort who had participated before in the Reykjavik Study.

**Atherosclerosis Risk in Communities Study (ARIC):** The ARIC study is a population-based prospective cohort study of cardiovascular disease sponsored by the National Heart, Lung, and Blood Institute (NHLBI). ARIC included 15,792 individuals, predominantly European American and African American, aged 45-64 years at baseline (1987-89), chosen by probability sampling from four US communities. Cohort members completed additional triennial follow-up examinations, a fifth exam in 2011-2013, a sixth exam in 2016-2017, and a seventh exam in 2018-2019, an eighth exam in 2020, a ninth exam in 2021-2022, and tenth exam in 2023, and an eleventh exam in 2024-2025. The twelfth exam is to be completed 2025-2026. The ARIC study has been described in detail previously (PMC8667593; Wright JD, Folsom AR, Coresh J, et al. The ARIC (Atherosclerosis Risk In Communities) Study: JACC Focus Seminar 3/8. J Am Coll Cardiol. 2021 Jun 15;77(23):2939-2959).

**ASPirin in Reducing Events in the Elderly (ASPREE):** The ASPREE study (n=19,114) is a randomized double-blind, placebo-controlled clinical trial, which aimed to determine whether daily 100-mg aspirin extended disability-free survival in healthy adults aged 70 years and older (65 years of age and older for U.S. minorities) with no history of diagnosed cardiovascular diseases, dementia, physical disability, or other life-threatening illness at enrolment. The design and protocol of the ASPREE trial have been reported previously [1-4]. All participants provided written informed consent. The study was approved by the Monash University and Alfred Hospital Human Research Ethics Committee (390/15) in Australia and site-specific Institutional Review Boards in the United States and registered on Clinicaltrials.gov (NCT01038583). Out of the 19,114 ASPREE trial participants, a total of 12,031 genotyped, unrelated participants (genetic relationship <0.05) aged ≥70 years with European ancestry (determined by genetic principal component analysis) were included in the CHARGE gene-lifestyle interaction studies.

**Bogalusa Heart Study (BHS):** The BHS study is population-based panel study to investigate the early natural history of cardiovascular disease and risk factors from childhood to adulthood among a biracial sample (65% white and 35% African American) of residents from Bogalusa, Louisiana. The study was established in 1973 by Dr. Gerald Berenson. To date, 9 surveys were conducted in children and adolescents aged 4 to 17 years, and 11 surveys were conducted among adults aged 18 to 51 years who were examined previously as children. At each survey, a standard questionnaire was used by trained research staffs to collect participants’ information on family status, levels of education, income, medical history and health behaviors. Clinical measures were collected following stringent protocols. The current study included 630 participants who were born between 1959 and 1979, examined at least once in childhood, and surveyed during the follow-up visit between 2013 and 2016. Genome-wide genotypes were assayed using the Illumina Human610 BeadChip. The genotype data were further imputed to the TOPMed reference panel using the TOPMed Imputation Server following a stringent genotype imputation protocol developed by the University of Michigan.

**Cameron County Hispanic Cohort (CCHC):** The CCHC (Cameron County Hispanic Cohort) is an ongoing, longitudinal study that was initiated in 2004 to investigate the burden of metabolic-related conditions and disparities in a Mexican American community. The ongoing study has recruited over 5000 participants through community-based research assistants who through a randomization process contact families living at the US-Mexico border in Cameron County, TX to voluntarily enroll in the program. They are then invited to visit the Clinical Research Unit located in facilities provided by Valley Baptist Medical Center, Brownsville, TX. Comprehensive clinical examinations and collection of specimens have been conducted and archived for further analysis. A subset of 4076 individuals have been genotyped using the Illumina MEGA array and the results have been imputed to TOPMed phase 8 reference data.

**The Cleveland Family Study (CFS):** is the largest family-based study of sleep apnea worldwide, consisting of 2,284 individuals (46% African American) from 361 families studied on up to 4 occasions over a period of 16 years. The study was begun in 1990 with the initial aims of quantifying the familial aggregation of sleep apnea. NIH renewals provided expansion of the original cohort (including increased minority recruitment) and longitudinal follow-up, with the last exam occurring in February 2006. 632 African Americans were genotyped on the Affymetrix array 6.0 platform through the CARe Consortium with suitable genotying quality control. A further 122 African-Americans had genotyping based on the Illumina OmniExpress + Exome platform. Genomes were imputed by TOPMed imputation server separately.

**Cardiovascular Health Study (CHS):** CHS is a population-based cohort study of risk factors for cardiovascular disease in adults 65 years of age or older conducted across four field centers (1). The original predominantly European ancestry cohort of 5,201 persons was recruited in 1989-1990 from random samples of the Medicare eligibility lists and an additional predominately African-American cohort of 687 persons was enrolled in 1992-93 for a total sample of 5,888. Blood samples were drawn from all participants at their baseline examination and DNA was subsequently extracted from available samples. European ancestry participants were excluded from the GWAS study sample due to prevalent coronary heart disease, congestive heart failure, peripheral vascular disease, valvular heart disease, stroke, or transient ischemic attack at baseline. After QC, genotyping was successful for 3271 European ancestry and 823 African-American participants. Phenotype data for this analysis were from the study visit in 1996-1997. CHS was approved by institutional review committees at each site and individuals in the present analysis gave informed consent including consent to use of genetic information for the study of cardiovascular disease.

1. Fried LP, Borhani NO, Enright P, Furberg CD, Gardin JM, Kronmal RA, et al. The Cardiovascular Health Study: design and rationale. Ann Epidemiol 1991; 1:263-76.

**Dose-Responses to Exercise Training (DR’s EXTRA):** The DR’s EXTRA study is a 4-year randomized controlled trial on the long-term effects of regular physical exercise and a healthy diet on atherosclerosis, cognition, and other health outcomes in a population-based random sample of Finnish men and women aged 55-74 years living in the city of Kuopio in 2002. The 3000 men and women who were invited to participate in the study were obtained from the national population registry. Altogether, 2062 men and women expressed their willingness to participate in the study, and 1479 of them attended the baseline measurements in 2005 - 2006. The prespecified exclusion criteria were medical or other conditions that prohibit engagement in the exercise intervention or the assessments, as judged by a physician. After these exclusions, 1410 individuals aged 57-78 years at baseline were randomized into the resistance exercise, aerobic exercise, diet, combined resistance exercise and diet, combined aerobic exercise and diet, or control group.

**European Prospective Investigation into Cancer and Nutrition (EPIC)-Norfolk:** The EPIC-Norfolk study (DOI 10.22025/2019.10.105.00004) is a prospective population-based cohort study which recruited 25,639 men and women aged 40-79 years at baseline between 1993 and 1997 from 35 participating general practices in Norfolk, UK [PMID:10466767]. Individuals attended for a baseline health check including the provision of blood samples for concurrent and future analysis. Further health check visits have been conducted since the baseline visit. Participants have contributed information about their diet, lifestyle and health through questionnaires and health checks over two decades. Sleep information was collected at the second heath check that was conducted in 1998-2000 (ages 42-82 years) with 15,786 participants. DNA has been extracted from all EPIC participants and stored blood has been analysed for an extensive range of classical and novel biomarkers. Sample quality control was performed including gender check, relatedness check, and ancestry check. The Norwich Local Research Ethics Committee granted ethical approval for the study. All participants gave written informed consent.

**The Fenland Study:** The Fenland study (DOI 10.22025/2017.10.101.00001) is a population-based cohort study that uses objective measures of disease exposure to investigate the influence of diet, lifestyle and genetic factors on the development of diabetes and obesity. The first phase of the Fenland Study was conducted between 2005 and 2015, and the volunteers are recruited from general practice lists in and around Cambridgeshire (Cambridge, Ely, and Wisbech) in the United Kingdom from birth cohorts from 1950–1975. Participants who attended the first phase of the study were invited to phase 2 of the study between 2014 and 2020, and the sleep data is available at this phase.

**Framingham Heart Study (FHS):** FHS began in 1948 with the recruitment of an original cohort of 5,209 men and women (mean age 44 years; 55 percent women). In 1971 a second generation of study participants was enrolled; this cohort (mean age 37 years; 52% women) consisted of 5,124 children and spouses of children of the original cohort. A third generation cohort of 4,095 children of offspring cohort participants (mean age 40 years; 53 percent women) was enrolled in 2002-2005 and are seen every 4 to 8 years. Details of study designs for the three cohorts are summarized elsewhere. At each clinic visit, a medical history was obtained with a focus on cardiovascular content, and participants underwent a physical examination including measurement of height and weight from which BMI was calculated.

**THL Biobank (FinnGen):** The THL Biobank is a Finnish biobank that has valuable research samples from all over Finland. It supports research investigating the causes of diseases, research on the impact of the hereditary, environmental and lifestyle factors to diseases. It hosts several significant population-based cohorts, disease-specific collections, and other nationally significant sample collections. For this study, datasets from 6 main studies contributed: DILGOM – samples from 2 time periods, Health 2017, FinnRisk 1992 – 2012 (a study with samples from 5 time points (1992, 1997, 2002, 2007 and 2012), GeneRISK, Health 2000 (samples from 2 periods, datasets called Health 2000 and Health 2011 and Kuusamo Health Examination Study 2011. Samples overlapping from studies which had multiple time points were excluded. For further information on the resource see https://thl.fi/en/research-and-development/thl-biobank/about-thl-biobank.

**Genetic and phenotypic determinants of blood pressure and other cardiovascular risk factors (GAPP):** GAPP is a population-based prospective cohort study involving a representative sample of initially healthy adults aged 25-41 years at baseline and residing in the Principality of Liechtenstein. Exclusion criteria were the presence of cardiovascular disease, diabetes, obstructive sleep apnea and a body mass index >35kg/m2. A standardized 12-lead ECG was obtained in all participants. Baseline characteristics were obtained in all participants using standard methodology. The main goal of GAPP is to assess the mechanisms for the development of cardiovascular risk factors over time.

**Genetic Epidemiology Network of Salt-Sensitivity (GenSalt)**: The GenSalt study is a unique NHLBI-sponsored family feeding-study designed to examine the interaction between genes and dietary sodium intake on BP. A detailed description of the GenSalt study design and participants has been reported previously^1^. Briefly, 3,142 participants from 633 Han families from rural, north China were ascertained through a proband with untreated pre-hypertension or stage-1 hypertension identified from a population-based BP screening. Individuals who had stage 2 hypertension, secondary hypertension, and a history of clinical cardiovascular disease or diabetes or were pregnant, heavy alcohol drinkers, or currently on a low-sodium diet or BP lowering medication were excluded from the study, with a total of 1,906 GenSalt probands and their siblings, spouses, and offspring eligible for the 7-day low sodium and 7-day high sodium dietary interventions. At baseline, a standard questionnaire was administered by a trained staff member to collect information on family pedigree, demographic characteristics, personal and family medical history, and lifestyle risk factors. Body weight and height were measured twice in light indoor clothing without shoes. Body mass index was calculated as weight in kilograms per height in square meters. BP was measured three times at the same time each morning during the three-day baseline examination by trained and certified. The mean of the 9 BP measures was used in subsequent analyses. Blood specimens were collected by venipuncture to measure lipids, creatinine, and other laboratory values. Among the 1,906 intervention participants, 1,881 underwent genome-wide genotyping and whole genome sequencing.

1. The GenSalt Collaborative Research Group. GenSalt: rationale, design, methods and baseline characteristics of study participants. *J Hyperten.*2007;21:639-646.
2. Perloff D, Grim C, Flack J, Frohglich ED, Hill M, McDonald M, Morgenstern BZ. Human blood pressure determination by sphygmomanometer. *Circulation.* 1993;88:2460–2470.

**GS (Generation Scotland):** Generation Scotland explores mental and physical health to improve the understanding, prevention and treatment of conditions for current and future generations. We are a collaboration between NHS Scotland, the University of Aberdeen, Dundee, Edinburgh and Glasgow, with over 40,000 participants across Scotland. Our mission is to advance mental and physical health research in areas including mental health, loneliness, cancer, dementia, reproductive health, the genetics of disease and much more. Data and samples have been collected from volunteers, with further information from linkage to medical records. Generation Scotland: Scottish Family Health Study (GS:SFHS)*, is a family-based study of about 24,000 volunteers across Scotland aged between 18-99 years. Ethical approval for the GS:SFHS study was obtained from the Tayside Committee on Medical Research Ethics (on behalf of the National Health Service).

*Smith, B. H. et al. Cohort Profile: Generation Scotland: Scottish Family Health Study (GS:SFHS). The study, its participants and their potential for genetic research on health and illness. Int J Epidemiol 42, 689–700 (2013).

**Hispanic Community Health Study/Study of Latinos (HCHS/SOL):** The HCHS/SOL is a multicenter prospective cohort of 16,000 Hispanic/Latino adults designed to investigate the role of acculturation in disease, and to identify other traits that impact Hispanic/Latino health. HCHS/SOL is the most diverse and comprehensive study of Hispanic/Latino health, with participants of Cuban, Puerto Rican, Dominican, Mexican or Central/South American origin. Participants were recruited through four sites affiliated with San Diego State University, Northwestern University in Chicago, Albert Einstein College of Medicine in Bronx, New York, and the University of Miami, using a census block and household sampling design. Study participants who were self-identified Hispanic/Latino and aged 18-74 years underwent extensive psycho-social, clinical assessments, and biospecimen collection during the baseline visit (2008-2011). A re-examination of the HCHS/SOL cohort was conducted during 2015-2017, and visit 3, which began in 2020, is ongoing. Annual telephone follow-up interviews have been conducted since study inception to determine health outcomes of interest. (dbGaP study accession number: phs000555).

**HCS (Hunter Community Study):** The HCS is a community-based longitudinal investigation that was commenced in Australia in 2004-2005. The study aims to investigate retired and near-retired persons by sampling older Australians aged 55–85, randomly selected from electoral rolls in a regional area on the heavily populated east coast (New South Wales). There were 3253 participants who completed at least some baseline measures. Follow-up was obtained through health record linkage up to 2017, representing over 10 years of health outcomes and hospitalizations.

**Insulin Resistance Atherosclerosis Study Family Study (IRASFS):** The IRASFS was a family study designed to examine the genetic and epidemiologic basis of glucose homeostasis traits and abdominal adiposity. Briefly, self-reported Mexican American pedigrees were recruited in San Antonio, TX and San Luis Valley, CO. Probands with large families were recruited from the initial non-family-based IRAS, which was modestly enriched for impaired glucose tolerance and T2D. Inclusion of IRASFS data is limited to 1040 normoglycemic individuals in 88 pedigrees with genotype data from the Illumina OmniExpress and Omni 1S arrays and imputation to the 1000 Genome Integrated Reference Panel (phase I).

**JHS (Jackson Heart Study):** The JHS is a longitudinal, community-based observational cohort study of 5,306 adults investigating the role of environmental and genetic factors in the development of cardiovascular disease in African Americans. Between 2000 and 2004, participants were recruited from a tri-county area (Hinds, Madison, and Rankin Counties) that encompasses Jackson, MS. Details of the design and recruitment for the Jackson Heart Study cohort has been previously published [1-3]. Briefly, approximately 30% of participants were former members of the Atherosclerosis Risk in Communities (ARIC) study. The remainder were recruited by either 1) random selection from the Accudata list, 2) commercial listing, 3) a constrained volunteer sample, in which recruitment was distributed among defined demographic cells in proportions designed to mirror those in the overall population, or through the Jackson Heart Study Family Study.

1. Wyatt SB, Diekelmann N, Henderson F, Andrew ME, Billingsley G, Felder SH, et al. A community-driven model of research participation: the Jackson Heart Study Participant Recruitment and Retention Study. *Ethn Dis*. 2003;13(4):438-55. PubMed PMID: 14632263.

2. Taylor HA, Jr., Wilson JG, Jones DW, Sarpong DF, Srinivasan A, Garrison RJ, et al. Toward resolution of cardiovascular health disparities in African Americans: design and methods of the Jackson Heart Study. *Ethn Dis*. 2005;15(4 Suppl 6):S6-4-17. PubMed PMID: 16320381.

3. Fuqua SR, Wyatt SB, Andrew ME, Sarpong DF, Henderson FR, Cunningham MF, et al. Recruiting African-American research participation in the Jackson Heart Study: methods, response rates, and sample description. *Ethn Dis*. 2005;15(4 Suppl 6):S6-18-29. PubMed PMID: 16317982.

**Korean Genome and Epidemiology Study (KOGES):** KoGES is a large-scale prospective cohort study with a comprehensive range of phenotypic measures and biological samples collected on approximately 210,000 individuals. KoGES’s long-term objective is to develop comprehensive and applicable healthcare guidelines for common complex diseases in Koreans, reduce the burden of chronic diseases and improve the quality of life. KoGES includes population-based cohorts, the community-based Ansan and Ansung study, the urban community-based health examinee study, and the rural community-based cardiovascular disease association study. The cohorts consist of community-dwellers and participants recruited from the national health examinee registry, men and women, aged ≥ 40 years at baseline. A total of 72,000 samples were genotyped with KoreanChip, a customized array optimized for the Korean population, and imputed using IMPUTE4 with 1000 Genomes Project Phase 3 data and the Korean reference genome as a reference panel. Measures of the baseline recruitment, and only genotyped samples that met the following exclusion criteria were used in the analyses: low call rate (<97%), excessive heterozygosity, excessive singletons, gender discrepancy, and cryptic first-degree relatives. SNPs with low HWE p value (<10^−6^) or low call rate (<95%) were excluded. Variants with imputation quality score (IQS) < 0.8 and MAF <1% were excluded after imputation.

1. Kim, Yeonjung, Bok-Ghee Han, and KoGES Group. "Cohort profile: the Korean genome and epidemiology study (KoGES) consortium." International journal of epidemiology 46.2 (2017): e20-e20.
2. Moon, Sanghoon, et al. "The Korea Biobank Array: design and identification of coding variants associated with blood biochemical traits." Scientific reports 9.1 (2019): 1-11.
3. Nam, Kisung, Jangho Kim, and Seunggeun Lee. "Genome-wide study on 72,298 individuals in Korean biobank data for 76 traits." Cell Genomics 2.10 (2022): 100189.

**Lothian Birth Cohort 1936 (LBC1936):** (https://lothian-birth-cohorts.ed.ac.uk/) LBC1936 consists of 1091 relatively healthy individuals born in 1936, most of whom took part in the Scottish Mental Survey of 1947 at the age of ~11 years old. They were recruited to a study to determine influences on cognitive ageing at age ~70 years, when almost all lived independently in the Lothian region of Scotland. For this project, data was drawn from LBC1936 Wave 3 (when sleep data was obtained). They have taken part in seven waves of testing in later life (at mean ages 70, 73, 76, 79, 82, 86 and 88 years). At each wave they underwent a series of cognitive and physical tests, biosampling, neuroimaging and other phenotyping.^1,2^

1. Deary IJ, Gow AJ, Pattie A, Starr JM. Cohort profile: the Lothian Birth Cohorts of 1921 and 1936. Int J Epidemiol 2012;41:1576-1584.

2. Taylor AM, Pattie A, Deary IJ. Cohort Profile Update: The Lothian Birth Cohorts of 1921 and 1936. Int J Epidemiol 2018;47:1042-1042r

**Lifelines**: (https://lifelines.nl/) Lifelines is a multi-disciplinary prospective population-based cohort study using a unique three-generation design to examine the health and health-related behaviors of 165,000 persons living in the North East region of The Netherlands. It employs a broad range of investigative procedures in assessing the biomedical, socio-demographic, behavioral, physical and psychological factors which contribute to the health and disease of the general population, with a special focus on multimorbidity. In addition, the Lifelines project comprises a number of cross-sectional sub-studies, which investigate specific age-related conditions. These include investigations into metabolic and hormonal diseases, including obesity, cardiovascular and renal diseases, pulmonary diseases and allergy, cognitive function and depression, and musculoskeletal conditions. All survey participants are between 18 and 90 years old at the time of enrollment. Recruitment has been going on since the end of 2006, and over 130,000 participants had been included by April 2013. At the baseline examination, the participants in the study were asked to fill in a questionnaire (on paper or online) before the first visit. During the first and second visit, the first or second part of the questionnaire, respectively, are checked for completeness, a number of investigations are conducted, and blood and urine samples are taken. Lifelines is a facility that is open for all researchers. Information on application and data access procedure is summarized on www.lifelines.nl. (Scholtens S, Smidt N, Swertz MA, Bakker SJ, Dotinga A, Vonk JM, et al. Cohort Profile: LifeLines, a three-generation cohort study and biobank. Int J Epidemiol. 2014 Dec 14.) Lifelines was genotyped in three stages using different chips: the first stage used the Illumina CytoSNP (CS) chip; the second the Illumina Global Screen Array (GSA); and the third the FinnGen Thermo Fisher Axiom from Affymetrix (Affy). Due to the differences in genotyping, these groups are analyzed separately. In case of close relatives being genotyped on different chips (as opposed to them being genotyped on the same chip, in which case the analysis software will account for their relatedness), the sample(s) on the less dense chip is excluded from the analysis; i.e. relatives on CS will always be excluded, those on GSA never.

**The Long Life Family Study (LLFS):** LLFS is a longitudinal, population-based multigenerational family cohort designed to study genetic, behavioral, and environmental factors in families exhibiting exceptional longevity. Families were sampled from four clinical centers: Boston University Medical Center in Boston, MA; Columbia College of Physicians and Surgeons in New York City, NY; the University of Pittsburgh in Pittsburgh, PA, USA; and the University of Southern Denmark, Denmark. The characteristics, recruitment, eligibility, and enrollment were previously described (PMID: 21258136, PMID: 34739053). The first clinical exam started in 2006 and recruited 4,953 individuals in 539 two-generational families that demonstrated clustering for exceptional survival in the upper generation. The second clinical exam (2014-2017) revisited 2,933 European descent individuals from 528 families. The third clinical exam (2021-) is recruiting the participants from second exam and a few new ones from the grandchild generation. The individuals were genotyped using ~2.3 million SNPs from the Illumina Omni chip, then imputed on Version R2 of the TOPMed reference panel using the Michigan Imputation Server (https://imputation.biodatacatalyst.nhlbi.nih.gov/#!), which used Eagle v2.4 for phasing and minimac4 v1.3.3 for imputation.

**Multi-Ethnic Study of Atherosclerosis (MESA):** The Multi-Ethnic Study of Atherosclerosis (MESA) is a study of the characteristics of subclinical cardiovascular disease and the risk factors that predict progression to clinically overt cardiovascular disease or progression of the subclinical disease. MESA consisted of a diverse, population-based sample of an initial 6,814 asymptomatic men and women aged 45-84. 38 percent of the recruited participants were white, 28 percent African American, 22 percent Hispanic, and 12 percent Asian, predominantly of Chinese descent. Participants were recruited from six field centers across the United States: Wake Forest University, Columbia University, Johns Hopkins University, University of Minnesota, Northwestern University and University of California - Los Angeles. Participants are being followed for identification and characterization of cardiovascular disease events, including acute myocardial infarction and other forms of coronary heart disease (CHD), stroke, and
congestive heart failure; for cardiovascular disease interventions; and for mortality. The first examination took place over two years, from July 2000 - July 2002. It was followed by five examination periods that were 17-20 months in length, including the recently completed Exam 6 (2016-2018). MESA Exam 7 will be completed by early 2024. Participants have been contacted every 9 to 12 months throughout the study to assess clinical morbidity and mortality. Informed consent was obtained for extensive data sharing (dbGaP) and genetic/omic studies, including candidate genes (NHLBI CARe), genome-wide scans (NHLBI SHARe), exome sequencing (NHBLI ESP) and, most recently, the NHLBI TOPMed program.

1. Bild DE, Bluemke DA, Burke GL, Detrano R, Diez Roux AV, Folsom AR, Greenland P, Jacob DR Jr, Kronmal R, Liu K, Nelson JC, O'Leary D, Saad MF, Shea S, Szklo M, Tracy RP. Multi-ethnic study of atherosclerosis: objectives and design. Am J Epidemiol. 2002 Nov 1;156(9):871-81. PubMed PMID: 12397006.

**The Netherlands Epidemiology of Obesity study (NEO):** The NEO was designed for extensive phenotyping to investigate pathways that lead to obesity-related diseases. The NEO study is a population-based, prospective cohort study that includes 6,671 individuals aged 45–65 years, with an oversampling of individuals with overweight or obesity. At baseline, information on demography, lifestyle, and medical history have been collected by questionnaires. In addition, samples of 24-h urine, fasting and postprandial blood plasma and serum, and DNA were collected. Genotyping was performed using the Illumina HumanCoreExome chip, which was subsequently imputed to the 1000 genome reference panel. Participants underwent an extensive physical examination, including anthropometry, electrocardiography, spirometry, and measurement of the carotid artery intima-media thickness by ultrasonography. In random subsamples of participants, magnetic resonance imaging of abdominal fat, pulse wave velocity of the aorta, heart, and brain, magnetic resonance spectroscopy of the liver, indirect calorimetry, dual energy X-ray absorptiometry, or accelerometry measurements were performed. The collection of data started in September 2008 and completed at the end of September 2012. Participants are currently being followed for the incidence of obesity-related diseases and mortality.

**Rotterdam Study**: The Rotterdam Study is a prospective, population-based cohort study among individuals living in the well-defined Ommoord district in the city of Rotterdam in The Netherlands. The aim of the study is to determine the occurrence of cardiovascular, neurological, ophthalmic, endocrine, hepatic, respiratory, and psychiatric diseases in elderly people. The cohort was initially defined in 1990 among approximately 7,900 persons, aged 55 years and older, who underwent a home interview and extensive physical examination at the baseline and during follow-up rounds every 3-4 years (RS-I). Cohort was extended in 2000/2001 (RS-II, 3,011 individuals aged 55 years and older) and 2006/2008 (RS-III, 3,932 subjects, aged 45 and older). In 2016, the most recent extension of the cohort was set up with 3005 persons aged 40 years and over, bringing the total Rotterdam Study cohort to 17,931 participants. Written informed consent was obtained from all participants and the Medical Ethics Committee of the Erasmus Medical Center, Rotterdam, approved the study.

**Study of Health in Pomerania (SHIP):** The Study of Health In Pomerania (SHIP) is a prospective longitudinal population-based cohort study in Mecklenburg-Western Pomerania assessing the prevalence and incidence of common diseases and their risk factors (PMID: 20167617 and 35348705). SHIP encompasses the two independent cohorts SHIP-START and SHIP-TREND. Participants aged 20 to 79 with German citizenship and principal residency in the study area were recruited from a random sample of residents living in the three local cities, 12 towns as well as 17 randomly selected smaller towns. Individuals were randomly selected stratified by age and sex in proportion to population size of the city, town or small towns, respectively. A total of 4,308 participants were recruited between 1997 and 2001 in the SHIP-START cohort. Between 2008 and 2012 a total of 4,420 participants were recruited in the SHIP-TREND cohort. Individuals were invited to the SHIP study center for a computer-assisted personal interviews and extensive physical examinations. The study protocol was approved by the medical ethics committee of the University of Greifswald. Oral and written informed consent was obtained from each of the study participants.

Genome-wide SNP-typing was performed using the Affymetrix Genome-Wide Human SNP Array 6.0 (SHIP-START samples), the Illumina Infinium HumanOmni2.5 BeadChip, or the Illumina Infinium Global Screening Array (SHIP-TREND samples). Array processing was carried out in accordance with the manufacturer’s standard recommendations. Genotypes were determined using the Birdseed2 clustering algorithm for SHIP-START, GenomeStudio Genotyping Module v1.0, and GenomeStudio 2.0 Genotyping Module (GenCall) for SHIP-TREND.

**UK Biobank (UKB):** UK Biobank (UKB, www.ukbiobank.ac.uk) is a large longitudinal biobank study in the United Kingdom which was established to improve understanding of the genetic and environmental causes of common diseases including cardiovascular diseases. In addition to self-reported disease outcomes and extensive health and life-style questionnaire data, UKB participants are being tracked through their NHS records and national registries (including cause of death and Hospital Episode Statistics). In 2017, UKB released the genotypes of 488,377 participants profiled with a custom SNP array. Genotyping QC was performed centrally by UKB, and genotypes imputed to Haplotype Reference Consortium (HRC) panel were released for 488,377 participants. Sample selection for the Gene-Lifestyle Interaction projects was based on available datasets for traits and lifestyle measures.

**Women’s Health Initiative (WHI):** is a long-term national health study that focuses on strategies for preventing common diseases such as heart disease, cancer and fracture in postmenopausal women. A total of 161,838 women aged 50–79 years old were recruited from 40 clinical centers in the US between 1993 and 1998(1, 2). WHI consists of an observational study, two clinical trials of postmenopausal hormone therapy (HT, estrogen alone or estrogen plus progestin), a calcium and vitamin D supplement trial, and a dietary modification trial. Study recruitment and exclusion criteria have been described previously(1, 2). Recruitment was done through mass mailing to age-eligible women obtained from voter registration, driver’s license and Health Care Financing Administration or other insurance list, with emphasis on recruitment of minorities and older women. Exclusions included participation in other randomized trials, predicted survival < 3 years, alcoholism, drug dependency, mental illness and dementia. For the CT, women were ineligible if they had a systolic BP > 200 mm Hg or diastolic BP > 105 mm Hg, a history of hypertriglyceridemia or breast cancer. Study protocols and consent forms were approved by the IRB at all participating institutions. Socio-demographic characteristics, lifestyle, medical history and self-reported medications were collected using standardized questionnaires at the screening visit. Genome wide association study (GWAS) non-overlapping samples are composed of (a) a case-control study (WHI Genomics and Randomized Trials Network – GARNET, which included all coronary heart disease, stroke, venous thromboembolic events and selected diabetes cases that happened during the active intervention phase in the WHI HT clinical trials and aged matched controls), (b) women selected to be "representative" of the HT trial (mostly younger white HT subjects that were also enrolled in the WHI memory study - WHIMS) and (c) the WHI SNP Health Association Resource (WHI SHARe), a randomly selected sample of 8,515 African American and 3,642 Hispanic women from WHI. Genotyping was performed using
Affymetrix 6.0 (WHI-SHARe), HumanOmniExpressExome-8v1_B (WHIMS) and Illumina HumanOmni1-Quad v1-0 B (GARNET). Quality control of the GWAS data included variant and sample call rates >95%, and included identification of genetically related individuals. Principal components were computed using methods developed by Price et al(3). Imputation was performed using the TOPMed Imputation Server and freeze 8 reference multi-ethnic panel (build hg38). Due to some overlap of WHI participants with the TOPMed reference panel, we re-calculated the estimated imputation quality based on only the samples not on TOPMed reference panel, to account for the over-estimation of imputation quality given by the imputation software(4). Variants with an imputation quality (Rsq) <0.3 were filtering out. After QC and exclusions from analysis protocol, the number of women included in analysis is 4,423 whites for GARNET, 5,202 white for WHIMS, 7,919 for SHARe African American and 3,377 for SHARe Hispanics. Analyses were performed using LinGxEScanR software.

1.	Design of the Women's Health Initiative clinical trial and observational study. The Women's Health Initiative Study Group. Control Clin Trials. 1998;19(1):61-109. Epub 1998/03/11. doi: S0197245697000780 [pii]. PubMed PMID: 9492970.

2.	Anderson GL, Manson J, Wallace R, Lund B, Hall D, Davis S, Shumaker S, Wang CY, Stein E, Prentice RL. Implementation of the Women's Health Initiative study design. Ann Epidemiol. 2003;13(9 Suppl):S5-17. Epub 2003/10/25. PubMed PMID: 14575938.

3.	Price AL, Patterson NJ, Plenge RM, Weinblatt ME, Shadick NA, Reich D. Principal components analysis corrects for stratification in genome-wide association studies. Nat Genet. 2006;38(8):904-9. doi: 10.1038/ng1847. PubMed PMID: 16862161.

4.	Sun Q, Liu W, Rosen JD, Huang L, Pace RG, Dang H, Gallins PJ, Blue EE, Ling H, Corvol H, Strug LJ, Bamshad MJ, Gibson RL, Pugh EW, Blackman SM, Cutting GR, O'Neal WK, Zhou YH, Wright FA, Knowles MR, Wen J, Li Y, Cystic Fibrosis Genome P. Leveraging TOPMed imputation server and constructing a cohort-specific imputation reference panel to enhance genotype imputation among cystic fibrosis patients. HGG Adv. 2022;3(2):100090. doi: 10.1016/j.xhgg.2022.100090. PubMed PMID: 35128485; PMCID: PMC8804187.

**The Cardiovascular Risk in Young Finns Study (YFS):** The YFS is a population-based follow up-study started in 1980. The main aim of the YFS is to determine the contribution made by childhood lifestyle, biological and psychological measures to the risk of cardiovascular diseases in adulthood. In 1980, over 3,500 children and adolescents all around Finland participated in the baseline study. The follow-up studies have been conducted mainly with 3-year intervals. The latest 30-year follow-up study was conducted in 2010-11 (ages 33-49 years) with 2,063 participants. The study was approved by the local ethics committees (University Hospitals of Helsinki, Turku, Tampere, Kuopio and Oulu) and was conducted following the guidelines of the Declaration of Helsinki. All participants gave their written informed consent.

#### Study Acknowledgments

**Age Gene/Environment Susceptibility Reykjavik Study (AGES):** This study has been funded by NIH contract N01-AG012100, HSSN271201200022C, the NIA Intramural Research Program, an Intramural Research Program Award (ZIAEY000401) from the National Eye Institute, an award from the National Institute on Deafness and Other Communication Disorders (NIDCD) Division of Scientific Programs (IAA Y2-DC_1004-02), Hjartavernd (the Icelandic Heart Association), and the Althingi (the Icelandic Parliament). The study is approved by the Icelandic National Bioethics Committee, VSN: 00-063. The researchers are indebted to the participants for their willingness to participate in the study.

**Atherosclerosis Risk in Communities Study (ARIC):** The ARIC study has been funded in whole or in part with Federal funds from the National Heart, Lung, and Blood Institute, National Institutes of Health, Department of Health and Human Services, under Contract nos. (75N92022D00001, 75N92022D00002, 75N92022D00003, 75N92022D00004, 75N92022D00005). The authors thank the staff and participants of the ARIC study for their important contributions. Funding was also supported by R01HL087641 and R01HL086694; National Human Genome Research Institute contract U01HG004402; and National Institutes of Health contract HHSN268200625226C. Infrastructure was partly supported by Grant Number UL1RR025005, a component of the National Institutes of Health and NIH Roadmap for Medical Research.

**ASPirin in Reducing Events in the Elderly (ASPREE):** The ASPREE study and Healthy Ageing Biobank were supported by an ASPREE Flagship cluster grant (including the Commonwealth Scientific and Industrial Research Organisation, Monash University, Menzies Research Institute, Australian National University, University of Melbourne); and grants (U01AG029824 and U19AG062682) from the National Institute on Aging and the National Cancer Institute at the National Institutes of Health, by grants (334047 and 1127060) from the National Health and Medical Research Council of Australia, and by Monash University and the Victorian Cancer Agency.

**Bogalusa Heart Study (BHS):** The BHS has been supported by multiple grants from the National Institute of Health, including R01AG077000, RF1AG041200, R01AG062309, and R33AG057983 from the National Institute on Aging, and R21HL161718 from the National Heart, Lung, and Blood Institute. The BHS study is extremely grateful to the participants for their willingness to participate in the study.

**Cameron County Hispanic Cohort (CCHC):** The CCHC is funded by the National Institutes of Health (NIH) CTSA UL1 TR00371, R01 HL142302-05A1, R01 DK127084-01A1, R01AG078452-01A1, CPRIT RP230063. The authors would like to acknowledge the invaluable contributions of the cohort staff, including Rocío Uribe, BSIE, and her team for their dedication to participant recruitment and data collection, as well as Marcela Morris, BS, for her assistance with laboratory procedures. We further extend our appreciation to the data management team, and to Norma Pérez-Olazarán, BBA, and Christina Villarreal, BA, for their administrative support. We are grateful to Valley Baptist Medical Center in Brownsville, Texas, for providing clinical space for the Center for Clinical and Translational Science Clinical Research Unit. Finally, we thank the community of Brownsville and the study participants for their generous participation and longstanding commitment to this research.

**The Cleveland Family Study (CFS):** was supported by grants from the National Institutes of Health (HL46380, M01 RR00080-39, T32-HL07567, RO1-46380).

**Cardiovascular Health Study (CHS):** This CHS research was supported by NHLBI contracts HHSN268201200036C, HHSN268200800007C, HHSN268201800001C, N01HC55222, N01HC85079, N01HC85080, N01HC85081, N01HC85082, N01HC85083, N01HC85086, 75N92021D00006; and NHLBI grants U01HL080295, R01HL085251, R01HL087652, R01HL105756, R01HL103612, R01HL120393, U01HL130114, and R01HL172803 with additional contribution from the National Institute of Neurological Disorders and Stroke (NINDS). Additional support was provided through R01AG023629 from the National Institute on Aging (NIA). A full list of principal CHS investigators and institutions can be found at CHS-NHLBI.org. The provision of genotyping data was supported in part by the National Center for Advancing Translational Sciences, CTSI grant UL1TR001881, and the National Institute of Diabetes and Digestive and Kidney Disease Diabetes Research Center (DRC) grant DK063491 to the Southern California Diabetes Endocrinology Research Center. The content is solely the responsibility of the authors and does not necessarily represent the official views of the National Institutes of Health.

**Dose-Responses to Exercise Training (DR’s EXTRA):** The DR's EXTRA Study was supported by the Ministry of Education and Culture of Finland (627, 722), the Academy of Finland (102318, 104943,123885, 211119), the European Commission FP6 Integrated Project EXGENESIS (LSHM-CT-2004-005272), the Research Committee of the Kuopio University Hospital Catchment Area (State Research Funding), Finnish Diabetes Association, Finnish Foundation for Cardiovascular Research, Päivikki and Sakari Sohlberg Foundation, Juho Vainio Foundation, the Social Insurance Institution of Finland (4/26/2010), and the city of Kuopio.

**European Prospective Investigation into Cancer and Nutrition (EPIC)-Norfolk:** The EPIC-Norfolk study (DOI 10.22025/2019.10.105.00004) has received funding from the Medical Research Council (MR/N003284/1 MC-UU_12015/1 and MC_UU_00006/1) and Cancer Research UK (C864/A14136). The genetics work in the EPIC-Norfolk study was funded by the Medical Research Council (MC_PC_13048). We are grateful to all the participants who have been part of the project and to the many members of the study teams at the University of Cambridge who have enabled this research.

**The Fenland Study:** The Fenland Study (DOI 10.22025/2017.10.101.00001) is funded by the Medical Research Council (MC_UU_12015/1). We are grateful to all the volunteers and to the General Practitioners and practice staff for assistance with recruitment. We thank the Fenland Study Investigators, Fenland Study Co-ordination team and the Epidemiology Field, Data and Laboratory teams. We further acknowledge support for genomics from the Medical Research Council (MC_PC_13046).

**Framingham Heart Study (FHS):** This research was conducted in part using data and resources from the Framingham Heart Study of the National Heart Lung and Blood Institute of the National Institutes of Health and Boston University School of Medicine. The analyses reflect intellectual input and resource development from the Framingham Heart Study investigators participating in the SNP Health Association Resource (SHARe) project. This work was partially supported by the National Heart, Lung and Blood Institute's Framingham Heart Study (Contract Nos. N01-HC-25195 and HSN268201500001I) and its contract with Affymetrix, Inc for genotyping services (Contract No. N02-HL-6-4278). This research was partially supported by grant R01DK122503 R01DK122503from the National Institute of Diabetes and Digestive and Kidney Diseases (MPIs: Kari North, Anne Justice, and Ching-Ti Liu).

**THL Biobank:** The data used for the research was obtained from THL Biobank (study number THLBB2022_64) https://www.thl.fi/biobank. We thank all study participants for their generous participation in THL Biobank. P.B.M. and S.F.S. acknowledge the support of the National Institute for Health and Care Research Barts Biomedical Research Centre (NIHR203330); a delivery partnership of Barts Health NHS Trust, Queen Mary University of London, St George’s University Hospitals NHS Foundation Trust and St George’s University of London. D.D. was funded by the NIH grant to DC as a subcontract. M.M.S recognizes his British Heart Foundation Clinical Research Training Fellowship (FS/CRTF/22/24353). We also acknowledge the Barts Charity grant G-002255 awarded to Dr M Sanghvi which contributed to access fees for this dataset. J.R. acknowledges fellowship RYC2021-031413-I from MCIN/AEI/10.13039/501100011033, and from the European Union ‘NextGenerationEU/PRTR’.

**Genetic and phenotypic determinants of blood pressure and other cardiovascular risk factors (GAPP):** The GAPP study was supported by the Liechtenstein Government, the Swiss Heart Foundation, the Swiss Society of Hypertension, the University of Basel, the University Hospital Basel, the Hanela Foundation, the Mach-Gaensslen Foundation, Schiller AG, and Novartis. We thank the GAPP staff and all GAPP study participants for their important contributions.

**Genetic Epidemiology Network of Salt-Sensitivity (GenSalt)**: This work was supported by a cooperative agreement project grant (U01HL072507, R01HL087263, and R01HL090682) from the National Heart, Lung and Blood Institute, National Institutes of Health, Bethesda, MD

**Generation Scotland (GS):** Generation Scotland received core support from the Chief Scientist Office of the Scottish Government Health Directorates [CZD/16/6] and the Scottish Funding Council [HR03006] and is currently supported by the Wellcome Trust [216767/Z/19/Z]. Genotyping of the GS:SFHS samples was carried out by the Genetics Core Laboratory at the Edinburgh Clinical Research Facility, University of Edinburgh, Scotland and was funded by the Medical Research Council UK and the Wellcome Trust (Wellcome Trust Strategic Award “STratifying Resilience and Depression Longitudinally” (STRADL) Reference 104036/Z/14/Z). C.H. was supported by an MRC programme grant “Quantitative Traits in Health and Disease (MC_UU_00007/10).

**The Hispanic Community Health Study/Study of Latinos (HCHS/SOL):** The HCHS/SOL is a collaborative study supported by contracts from the National Heart, Lung, and Blood Institute (NHLBI) to the University of North Carolina (HHSN268201300001I / N01-HC-65233), University of Miami (HHSN268201300004I / N01-HC-65234), Albert Einstein College of Medicine (HHSN268201300002I / N01-HC-65235), University of Illinois at Chicago (HHSN268201300003I / N01- HC-65236 Northwestern Univ), and San Diego State University (HHSN268201300005I / N01-HC-65237). The following Institutes/Centers/Offices have contributed to the HCHS/SOL through a transfer of funds to the NHLBI: National Institute on Minority Health and Health Disparities, National Institute on Deafness and Other Communication Disorders, National Institute of Dental and Craniofacial Research, National Institute of Diabetes and Digestive and Kidney Diseases, National Institute of Neurological Disorders and Stroke, NIH Institution-Office of Dietary Supplements. The Genetic Analysis Center at the University of Washington was supported by NHLBI and NIDCR contracts (HHSN268201300005C AM03 and MOD03).

**Hunter Community Study (HCS):** The authors would like to thank the men and women participating in the HCS as well as all the staff, investigators and collaborators who have supported or been involved in the project to date. The authors would also like to thank the Hunter Medical Research Institute who provided media support during the initial recruitment of participants; and Dr Anne Crotty, Prof. Rodney Scott and Associate Prof. Levi who provided financial support towards freezing costs for the long-term storage of participant blood samples.

**Insulin Resistance Atherosclerosis Study Family Study (IRASFS):** The IRASFS was supported by the National Heart Lung and Blood Institute (HL060944, HL061019, and HL060919). Genotyping and analysis for this study was supported by the GUARDIAN Consortium with grant support from the National Institute of Diabetes, Digestive, and Kidney Diseases (NIDDK; DK085175 and DK118062) and in part by UL1TR000124 (CTSI) and DK063491 (DRC). The authors thank study investigators, staff, and participants for their valuable contributions.

**Integrated Methods of Analysis for Genetic Epidemiology (IMAGE):** A program project grant (PO1-CA196569) that supported the development of statistical methods and the LinGxEScanR software used in this paper.

**Jackson Heart Study (JHS):** The Jackson Heart Study is supported by Contracts HHSN268201800010I, HHSN268201800011I, HHSN268201800012I, HHSN268201800013I, HHSN268201800014I, HHSN268201800015I from the National Heart Lung and Blood Institute (NHLBI) with additional support from the National Institute of Minority Health and Health Disparities (NIMHD). The authors also wish to thank the staffs and participants of the JHS. The views expressed in this manuscript are those of the authors and do not necessarily represent the views of the National Heart, Lung, and Blood Institute; the National Institute of Minority Health and Health Disparities (NIMHD); the National Institutes of Health; or the U.S. Department of Health and Human Services.

**Korean Genome and Epidemiology Study (KOGES):** Data were collected and supported by the National Research Institute of Health, Centers for Disease Control and Prevention, and Ministry for Health and Welfare, Republic of Korea. The authors thank the staff and participants of the KoGES study for their important contributions. Additional support was provided through grants the Brain Pool Plus (BP+, Brain Pool+) Program (2020H1D3A2A03100666) and Sejong Science Fellowship (2022R1C1C2006474) through the National Research Foundation of Korea (NRF) funded by the Ministry of Science and ICT.

**Lothian Birth Cohort 1936 (LBC1936):** The authors thank all LBC1936 study participants and research team members who have contributed, and continue to contribute, to ongoing studies. LBC1936 is supported by the Biotechnology and Biological Sciences Research Council (BBSRC), and the Economic and Social Research Council [BB/W008793/1] (which supports S.E.H.), Age UK (Disconnected Mind project), the Milton Damerel Trust, and the University of Edinburgh. SRC is supported by a Sir Henry Dale Fellowship jointly funded by the Wellcome Trust and the Royal Society (221890/Z/20/Z). S.E.H. is supported by a National Institutes of Health (NIH) research grant (U01AG083829). Genotyping was funded by the BBSRC (BB/F019394/1).

**Lifelines Cohort Study:**

Raul Aguirre-Gamboa (1), Patrick Deelen (1), Lude Franke (1), Jan A Kuivenhoven (2), Esteban A Lopera Maya (1), Ilja M Nolte (3), Serena Sanna (1), Harold Snieder (3), Morris A Swertz (1), Peter M. Visscher (3,4), Judith M Vonk (3), Cisca Wijmenga (1), Naomi Wray (4)

(1) *Department of Genetics, University of Groningen, University Medical Center Groningen, The Netherlands*
(2) *Department of Pediatrics, University of Groningen, University Medical Center Groningen, The Netherlands*
(3) *Department of Epidemiology, University of Groningen, University Medical Center Groningen, The Netherlands*
(4) *Institute for Molecular Bioscience, The University of Queensland, Brisbane, Queensland, Australia.*

The Lifelines Biobank initiative has been made possible by funding from the Dutch Ministry of Health, Welfare and Sport, the Dutch Ministry of Economic Affairs, the University Medical Center Groningen (UMCG the Netherlands), University of Groningen and the Northern Provinces of the Netherlands. The generation and management of GWAS genotype data for the Lifelines Cohort Study is supported by the UMCG Genetics Lifelines Initiative (UGLI). UGLI is partly supported by a Spinoza Grant from NWO, awarded to Cisca Wijmenga.

The authors wish to acknowledge the services of the Lifelines Cohort Study, the contributing research centers delivering data to Lifelines, and all the study participants.

**The Long Life Family Study (LLFS):** This work was supported by the National Institute on Aging (U01AG023746, U01AG023712, U01AG023749, U01AG023755, U01AG023744, and U19AG063893).

**Multi-Ethnic Study of Atherosclerosis (MESA):** MESA and the MESA SHARe project are conducted and supported by the National Heart, Lung, and Blood Institute (NHLBI) in collaboration with MESA investigators. Support for MESA is provided by contracts 75N92025D00022, 75N92020D00001, HHSN268201500003I, N01-HC-95159, 75N92025D00026, 75N92020D00005, N01-HC-95160, 75N92020D00002, N01-HC-95161, 75N92025D00024, 75N92020D00003, N01-HC-95162, 75N92025D00027, 75N92020D00006, N01-HC-95163, 75N92025D00025, 75N92020D00004, N01-HC-95164, 75N92025D00028, 75N92020D00007, N01-HC-95165, N01-HC-95166, N01-HC-95167, N01-HC-95168, N01-HC-95169, UL1-TR-000040, UL1-TR-001079, UL1-TR-001420, UL1TR001881, DK063491, and R01HL105756. The authors thank the MESA participants and the MESA investigators and staff for their valuable contributions. A full list of participating MESA investigators and institutions can be found at http://www.mesa-nhlbi.org.

**The Netherlands Epidemiology of Obesity study (NEO):** The authors of the NEO study thank all individuals who participated in the Netherlands Epidemiology in Obesity study, all participating general practitioners for inviting eligible participants and all research nurses for collection of the data. We thank the NEO study group, Petra Noordijk, Pat van Beelen and Ingeborg de Jonge for the coordination, lab and data management of the NEO study. The genotyping in the NEO study was supported by the Centre National de Génotypage (Paris, France), headed by Jean-Francois Deleuze. The NEO study is supported by the participating Departments, the Division and the Board of Directors of the Leiden University Medical Center, and by the Leiden University, Research Profile Area Vascular and Regenerative Medicine.

**RS (Rotterdam Study):** The Rotterdam Study is funded by Erasmus Medical Center and Erasmus University, Rotterdam, Netherlands Organization for the Health Research and Development (ZonMw), the Research Institute for Diseases in the Elderly (RIDE), the Ministry of Education, Culture and Science, the Ministry for Health, Welfare and Sports, the European Commission (DG XII), and the Municipality of Rotterdam. The authors are grateful to the study participants, the staff from the Rotterdam Study and the participating general practitioners and pharmacists. The generation and management of GWAS genotype data for the Rotterdam Study was executed by the Human Genotyping Facility of the Genetic Laboratory of the Department of Internal Medicine, Erasmus MC, Rotterdam, The Netherlands. The GWAS datasets are supported by the Netherlands Organisation of Scientific Research NWO Investments (nr. 175.010.2005.011, 911-03-012), the Genetic Laboratory of the Department of Internal Medicine, Erasmus MC, the Research Institute for Diseases in the Elderly (014-93-015; RIDE2), the Netherlands Genomics Initiative (NGI)/Netherlands Organisation for Scientific Research (NWO) Netherlands Consortium for Healthy Aging (NCHA), project nr. 050-060-810. We thank Pascal Arp, Mila Jhamai, Marijn Verkerk, Lizbeth Herrera, Marjolein Peters and Carolina Medina-Gomez for their help in creating the GWAS database, and Karol Estrada, Yurii Aulchenko and Carolina Medina-Gomez for the creation and analysis of imputed data.

**Study of Health in Pomerania (SHIP):** SHIP is part of the Community Medicine Research net of the University of Greifswald, Germany, which is funded by the Federal Ministry of Education and Research (grants no. 01ZZ9603, 01ZZ0103, and 01ZZ0403), the Ministry of Cultural Affairs as well as the Social Ministry of the Federal State of Mecklenburg-West Pomerania, and the network ‘Greifswald Approach to Individualized Medicine (GANI_MED)’ funded by the Federal Ministry of Education and Research (grant 03IS2061A). Genome-wide data were supported by the Federal Ministry of Education and Research (grant no. 03ZIK012) and a joint grant from Siemens Healthcare, Erlangen, Germany and the Federal State of Mecklenburg- West Pomerania.

**UK Biobank (UKB):** This research has been conducted using the UK Biobank Resource under Application Number 8343. This research used data assets made available by National Safe Haven as part of the Data and Connectivity National Core Study, led by Health Data Research UK in partnership with the Office for National Statistics and funded by UK Research and Innovation (grant ref MC_PC_20029). Copyright © (2022), NHS Digital.  Re-used with the permission of the NHS Digital [and/or UK Biobank].  All rights reserved.

**Women’s Health Initiative (WHI):** The WHI program is funded by the National Heart, Lung, and Blood Institute, National Institutes of Health, U.S. Department of Health and Human Services through contracts HHSN268201600018C, HHSN268201600001C, HHSN268201600002C, HHSN268201600003C, and HHSN268201600004C. The authors thank the WHI investigators and staff for their dedication, and the study participants for making the program possible. A full listing of WHI investigators can be found at: https://www-whi-org.s3.us-west-2.amazonaws.com/wp-content/uploads/WHI-Investigator-Long-List.pdf. Additional support to NF was provided by NIH DK117445 and MD012765.

**The Cardiovascular Risk in Young Finns Study (YFS):** The Young Finns Study has been financially supported by the Academy of Finland: grants 356405, 322098, 286284, 134309 (Eye), 126925, 121584, 124282, 129378 (Salve), 117797 (Gendi), 141071 (Skidi), 349708, 330809, and 338395; the Social Insurance Institution of Finland; Competitive State Research Financing of the Expert Responsibility area of Kuopio, Tampere and Turku University Hospitals (grant X51001); Juho Vainio Foundation; Paavo Nurmi Foundation; Finnish Foundation for Cardiovascular Research ; Finnish Cultural Foundation; The Sigrid Juselius Foundation; Tampere Tuberculosis Foundation; Emil Aaltonen Foundation; Yrjö Jahnsson Foundation; Signe and Ane Gyllenberg Foundation; Diabetes Research Foundation of Finnish Diabetes Association; EU Horizon 2020 (grant 755320 for TAXINOMISIS and grant 848146 for To Aition); European Research Council (grant 742927 for MULTIEPIGEN project); Tampere University Hospital Supporting Foundation, Finnish Society of Clinical Chemistry, the Cancer Foundation Finland; pBETTER4U_EU (Preventing obesity through Biologically and bEhaviorally Tailored inTERventions for you, project number: 101080117); CVDLink (EU grant nro. 101137278), and the Jane and Aatos Erkko Foundation.
